# COVID-19 as cause of viral sepsis: A Systematic Review and Meta-Analysis

**DOI:** 10.1101/2020.12.02.20242354

**Authors:** Eleni Karakike, Evangelos J. Giamarellos-Bourboulis, Miltiades Kyprianou, Carolin Fleischmann-Struzek, Mathias W. Pletz, Mihai G. Netea, Konrad Reinhart, Evdoxia Kyriazopoulou

**Author notes:** equal contribution (Konrad Reinhart, Evdoxia Kyriazopoulou). **Corresponding Author:** Evangelos J. Giamarellos-Bourboulis, MD, PhD, 4^th^ Department of Internal Medicine, ATTIKON University General Hospital, 1 Rimini Street 124 62 Athens, Greece.

## Abstract

**Importance:** COVID-19 is a heterogenous disease most frequently causing respiratory tract infection but in its severe forms, respiratory failure and multiple organ dysfunction syndrome may occur, resembling sepsis. The prevalence of viral sepsis among COVID-19 patients is still unclear.

**Objective:** We aimed to describe this in a systematic review.

**Data sources:** MEDLINE(PubMed), Cochrane and Google Scholar databases were searched for studies reporting on patients hospitalized with confirmed COVID-19, diagnosed with sepsis or infection-related organ dysfunctions or receiving organ replacement therapy.

**Study selection:** Eligible were full-text English articles of randomized and non-randomized clinical trials and observational studies reporting on patients with confirmed COVID-19, who are diagnosed with sepsis or have infection-related organ dysfunctions. Systematic reviews, editorials, conference abstracts, animal studies, case reports, articles neither in English nor full-text, and studies with fewer than 30 participants were excluded.

**Data extraction and synthesis:** All eligible studies were included in a narrative synthesis of results and after reviewing all included studies a meta-analysis was conducted. Separate sensitivity analyses were conducted per adult vs pediatric populations and per Intensive Care Unit (ICU) vs non-ICU populations.

**Main outcomes and measures:** Primary endpoint was the prevalence of sepsis using Sepsis-3 criteria among patients with COVID-19 and among secondary, new onset of infection-related organ dysfunction. Outcomes were expressed as proportions with respective 95% confidence interval (CI).

**Results:** Of 1,903 articles, 104 were analyzed. The prevalence of sepsis in COVID-19 was 39.9% (95% CI, 35.9-44.1; I^2^, 99%). In sensitivity analysis, sepsis was present in 25.1% (95% CI, 21.8-28.9; I^2^ 99%) of adult patients hospitalized in non-Intensive-Care-Unit (ICU) wards (40 studies) and in 83.8 (95% CI, 78.1-88.2; I^2^,91%) of adult patients hospitalized in the ICU (31 studies). Sepsis in children hospitalized with COVID-19 was as high as 7.8% (95% CI, 0.4-64.9; I^2^, 97%). Acute Respiratory Distress Syndrome was the most common organ dysfunction in adult patients in non-ICU (27.6; 95% CI, 21.6-34.5; I^2^, 99%) and ICU (88.3%; 95% CI, 79.7-93.5; I^2^, 97%)

**Conclusions and relevance:** Despite the high heterogeneity in reported results, sepsis frequently complicates COVID-19 among hospitalized patients and is significantly higher among those in the ICU. PROSPERO registration number: CRD42020202018. No funding.

**KEY POINTS:** *Question:* What is the prevalence of viral sepsis by Sepsis-3 definition among hospitalized patients with COVID-19?

*Findings:* In this systematic review and meta-analysis, we systematically reviewed published literature for evidence of organ failure in COVID-19, to estimate the prevalence of viral sepsis in this setting, by means of SOFA score calculation. The prevalence of sepsis in COVID-19 was 39.9% (95% CI, 35.9-44.1; I^2^, 99%).

*Meaning:* This is the first study to address the burden of viral sepsis in hospitalized patients with COVID-19, a highly heterogenous infection ranging from asymptomatic cases to severe disease leading to death, as reflected in the high heterogeneity of this study.

## INTRODUCTION

COVID-19, caused by the severe acute respiratory syndrome coronavirus (SARS-CoV)-2, is recognized as a pandemic, affecting 230 countries with 54,771,888 confirmed cases and 1,324,249 deaths worldwide, as of 17 November 2020 (1). Several patients are complicated by respiratory insufficiency and they are in need for mechanical ventilation (2). First reports of patients in Wuhan described in addition of lung disease the presence of other organ failures including acute kidney injury, liver dysfunction, coagulopathy, confusion and shock; such an involvement resembles the systemic counterparts of bacterial and viral sepsis (3-5). The current Sepsis-3 definitions define sepsis as a life-threatening organ dysfunction due to the dysregulated host response to an infection. The same definitions introduce the Sequential Organ Failure Assessment (SOFA) score as a measure of organ dysfunction (6). According to World Health Organization (WHO), manifestations of sepsis and septic shock can be also the fatal frequent pathway of infections with highly transmissible pathogens of public health concern such as avian and swine influenza viruses, as well as corona viruses (7). Severe respiratory failure of COVID-19 is accompanied by complex immune dysregulation of the host (8). As a consequence, all elements of the new Sepsis-3 definition may apply for COVID-19 i.e. life-threatening organ dysfunction, dysregulated host response, and viral infection (9).

With this in mind, we systematically reviewed published evidence for COVID-19 as a cause of viral sepsis using the SOFA score as a classification tool. In parallel the prevalence of different organ dysfunctions, the need for ICU admission, the association of sepsis presence with mortality and the presence of alterations of inflammation and coagulation markers were analyzed.

## METHODS

### Search strategy and selection criteria

The literature search and review process were conducted according to the Preferred Reporting Items for Systematic reviews and Meta-Analyses (PRISMA) statement (10), based on a pre-specified protocol (PROSPERO database registration number: CRD42020202018).

The search of the peer-reviewed medical literature was conducted across MEDLINE (PubMed), Cochrane and Google Scholar databases using the following terms: “COVID-19” or “SARS-CoV-2” and “sepsis”, “organ failure”, “organ dysfunction”. Detailed search strategy is provided in Supplement. Inclusion criteria comprised randomized and non-randomized clinical trials and observational studies reporting on the proportion of patients diagnosed with confirmed COVID-19, who were diagnosed with sepsis, had infection-related organ dysfunctions, or received organ replacement therapy (dialysis, mechanical ventilation, extracorporeal membrane oxygenation [ECMO], liver replacement therapy). Sepsis was defined i) as sepsis according to Sepsis-3 definitions (i.e. any at least two-point SOFA score at presentation or any at least two-point-increase of the baseline SOFA score during hospitalization; ii) as severe sepsis according to Sepsis 1/2 criteria; or iii) by relevant ICD codes (7,11,12). Only articles published as full text in English were included, while systematic reviews, editorials, conference abstracts, animal studies, case reports, articles not written in English or not providing full-text, and studies with fewer than 30 participants were excluded.

The literature search was conducted on August 27, 2020 and repeated on October 3, 2020 by two independent authors (E.Ky and E.Ka). The same reviewers assessed all articles by title, abstract and complete text to find those meeting the predefined eligibility criteria and extracted information as follows: first author name, country of origin, publication year and month, study design, total number of patients reported, criteria for enrollment in the study, number of patients presenting sepsis, new onset organ dysfunction, organ support/replacement therapy, number of patients requiring ICU treatment, ICU discharge, hospital discharge and 30-day mortality. Any controversies were resolved by a third reviewer (E.J.G.B). All authors of the selected articles were contacted to provide relevant data.

Each study was evaluated by both reviewers in terms of quality of the provided data with the Methodological index for non-randomized studies (MINORS) (13). An additional assessment of certainty in extraction of the primary endpoint was made, rated by means of a four-star scale, from zero (one star) to high uncertainty (four stars), in order to assess causes for potential heterogeneity. Articles providing the exact number of patients fulfilling Sepsis-3 criteria (either in the original publication or after contacting corresponding authors), were qualified as zero uncertainty; articles reporting mean/ median SOFA score with respective standard deviation (SD)/ interquartile range (IQR) were qualified as low uncertainty, whereas articles allowing extraction of SOFA score ≥ 2, based at least on one reported specific organ dysfunction were characterized as intermediate and articles where only severity of disease (e.g. need for mechanical ventilation, need for ICU, or presence of “critical illness”) could be used as a proxy for sepsis were considered of high uncertainty. The following assumptions were made: mechanical ventilation and non-invasive mechanical ventilation were considered as a PaO2: FiO2 ratio< 300, and thus as SOFA score of ≥2. Among articles allowing extraction of SOFA score ≥2 for different organ dysfunctions, a conservative approach for calculating the primary outcome was followed and only one organ was considered (the one with the maximum number of affected patients within the cohort). Among articles reporting medians (interquartile range), outcomes were calculated as the minimum *n* observed. In order to avoid including patients with sepsis due to secondary bacterial infections, corresponding authors of publications reporting “sepsis” among outcomes were contacted for clarification.

### Endpoints and outcome measures

The primary endpoint was the prevalence of sepsis among COVID-19 patients. The outcome measure was the proportion of patients with COVID-19 who fulfill the criteria for sepsis with 95% confidence intervals (CI). Secondary outcomes included: i) the prevalence of new onset infection-related organ dysfunction; ii) the prevalence of organ support and/or replacement (invasive and non-invasive mechanical ventilation, including high-flow nasal cannula, vasopressors, ECMO, liver and renal replacement therapy); iii) the prevalence of ICU admission; iv) the mortality of COVID-19 patients with and without sepsis; and iv) the presence of alterations of inflammation and coagulation markers. The outcome measure for each secondary outcome was the respective proportion of patients with COVID-19 and 95% CI.

### Sensitivity analysis

Separate sensitivity analyses were planned per adult vs pediatric populations and per ICU vs non-ICU populations. Also, sensitivity analysis was performed among studies with low and zero vs high uncertainty regarding the primary outcome. An additional sensitivity analysis was planned for studies reporting patients without selection criteria (all-comers) vs those reporting on specific selected groups. Articles reporting on mixed cohorts (both ICU and non-ICU or adult and pediatric) were included in the primary analysis as a whole and in the sensitivity analyses with the subgroup concerned in the denominator.

### Statistical analysis

All eligible studies were included in a narrative synthesis and after reviewing all included studies. Meta-analysis was performed using the R software v. 4.0.2 (14) after installing the packages “meta” (15), “metaphor” (16) and “dmetar” (17). Some results were validated in the Review Manager (RevMan, The Cochrane Collaboration) Version 5.4. In all cases the random effects model was employed. For each analysis, the corresponding forest plot was produced, while publication bias was assessed with the Egger’s test via funnel plot asymmetry (18).

## RESULTS

The literature search yielded 1,903 articles; of these, after removal of duplicates and of articles with irrelevant title, 238 were screened full-text by the reviewers (E. Ky and E. Ka). After removal of articles fulfilling exclusion criteria, 104 articles including a total of 157,063 patients were finally analyzed (Figure 1). Data on primary endpoint were provided in 92 studies including a total of 147,881 patients. Of the 104 included studies, mainly observational retrospective, 42 were reporting results from China (19-60), 19 from the U.S.A (61-79), 5 from the U.K (80-84), 24 across Europe, mainly Italy, Spain, France and the Netherlands (85-109), 10 from other countries (110-119) and 3 were international (120-122). All studies reported data on hospitalized patients due to COVID-19 and 35 of them included data for ICU nnnnnn(20, 25, 33, 36, 37, 41, 44, 45, 48, 52, 53, 56, 58, 66, 72, 73, 81-84, 86-89, 95, 96, 98, 101, 103, 106, 107, 109, 111, 119). Eight studies were pediatric (59, 60, 78, 79, 83, 84, 118, 119). The majority of articles included patients without any selection criteria (all-comers), whereas forty-five focused on specific subgroups, such as critically ill, cancer or rheumatologic patients, patients on mechanical ventilation etc. No studies were found that reported sepsis based on Sepsis-3 or Sepsis-1/2 criteria per se in the original publication; zero uncertainty was only achieved when corresponding authors contacted, provided the exact number of patients presenting sepsis based on sepsis-3. Characteristics of the included studies are presented in eTable 1 and quality assessment with MINORS of each in eTable 2.

**Figure 1.**
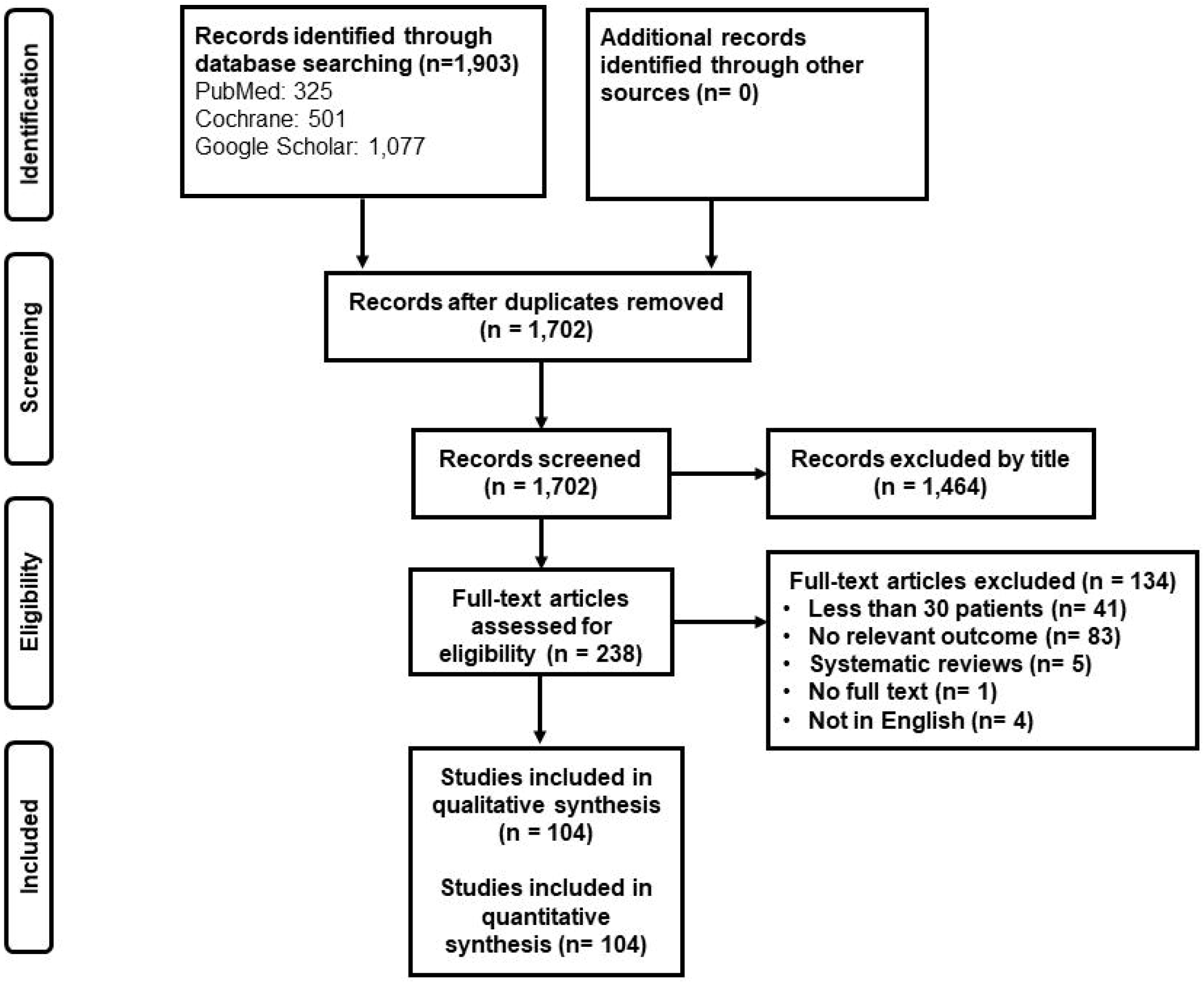
Study selection.

### Primary endpoint

The pooled estimate for the prevalence of sepsis among COVID-19 patients was 39.9% (95% CI, 35.9-44.1; I^2^, 99%) as assessed from 92 studies reporting data on the primary endpoint (eFigure 1). Due to the high heterogeneity observed, the results of sensitivity analyses are presented in Figure 2. In sensitivity analysis focusing on ICU versus non-ICU cohorts, among forty studies of a total of 112,796 patients, the pooled estimate for sepsis prevalence was 25.1% (95% CI, 21.8-28.9; I^2^ 99%) in the general ward (Figure 2A). The respective estimate for adult patients in the ICU was 83.8% (95% CI, 78.1-88.2; I^2^ 91%) among 31 articles assessed including 3,528 patients (p<0.0001) (Figure 2B). In sensitivity analysis among pediatric cohorts, sepsis in children hospitalized outside the ICU was as high as 7.8% (95% CI, 0.4-64.9; I^2^, 97%) among 3 studies assessed (Figure 2C), whereas only one study provided data for ICU (all 78 children septic) (84). Respective funnel plots of the above analyses are provided in eFigure 2. Sensitivity analyses including only articles with zero uncertainty (4 studies outside the ICU and 3 in the ICU, eFigure 3) and high uncertainty (13 studies outside the ICU and 3 in the ICU, eFigure 4) provided similar pooled estimates of sepsis prevalence outside of the ICU, which were 17.1% (95%CI, 6.4-38.4; I^2^, 97%) in zero uncertainty analysis and 21.9% (95%CI, 16.9-27.9; I^2^, 93%) in high uncertainty analysis. However, the pooled estimate in the ICU was 95.7% (95%CI, 75.5-99.4; I^2^, 65%) and 34.7% (95%CI, 9.9-72.0, I^2^, 92%) in zero and high uncertainty analysis respectively (p<0.0001), reducing the heterogeneity and yielding a significant difference between the two groups.

**Figure 2.**
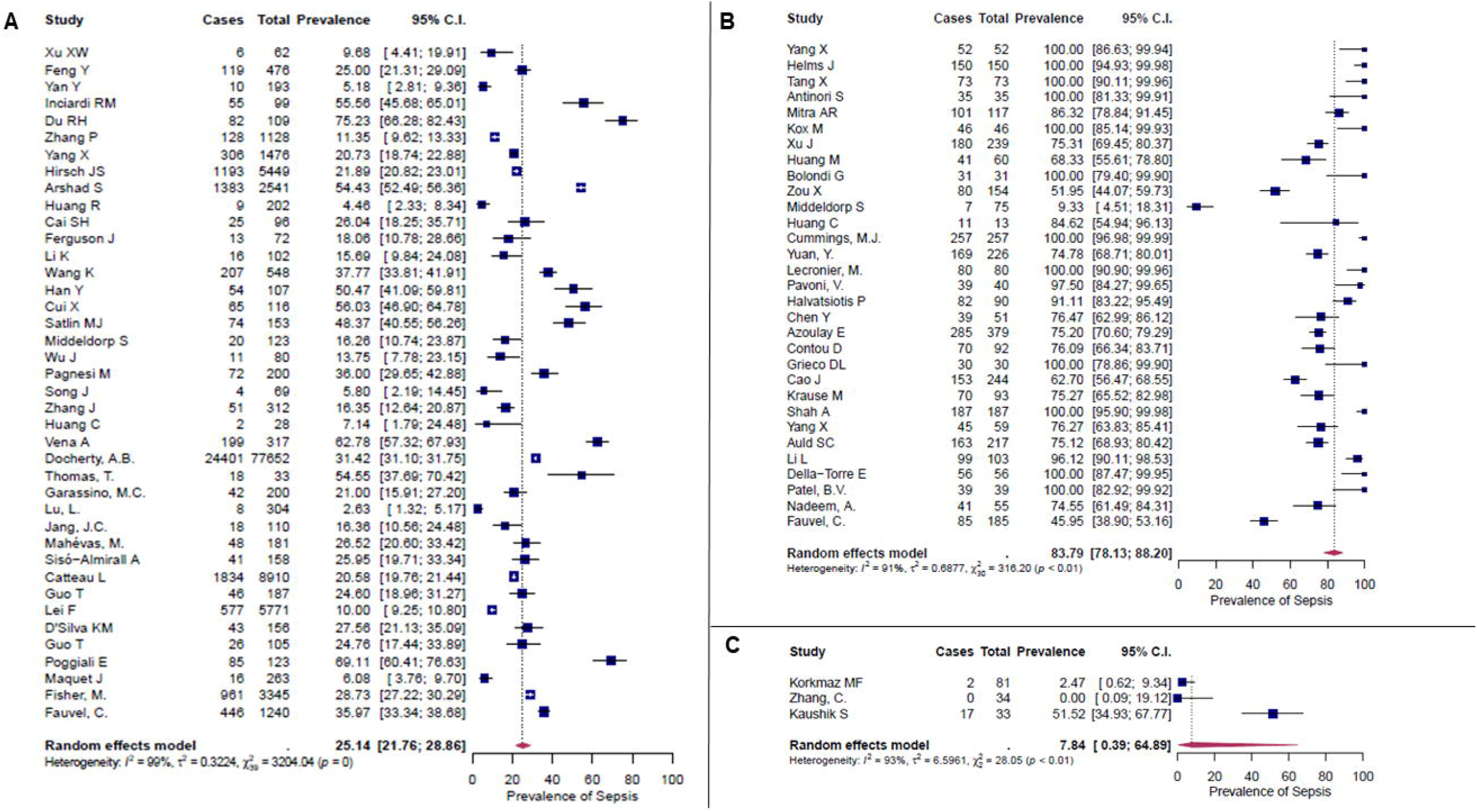
Forest plot of sepsis prevalence in A) adult patients outside the Intensive Care Unit (ICU); B) adult patients in the ICU; and C) pediatric patients outside the ICU. Only studies with zero and low uncertainty have been included. All included studies have been published in 2020. Abbreviation: CI, confidence interval.

**Figure 3.**
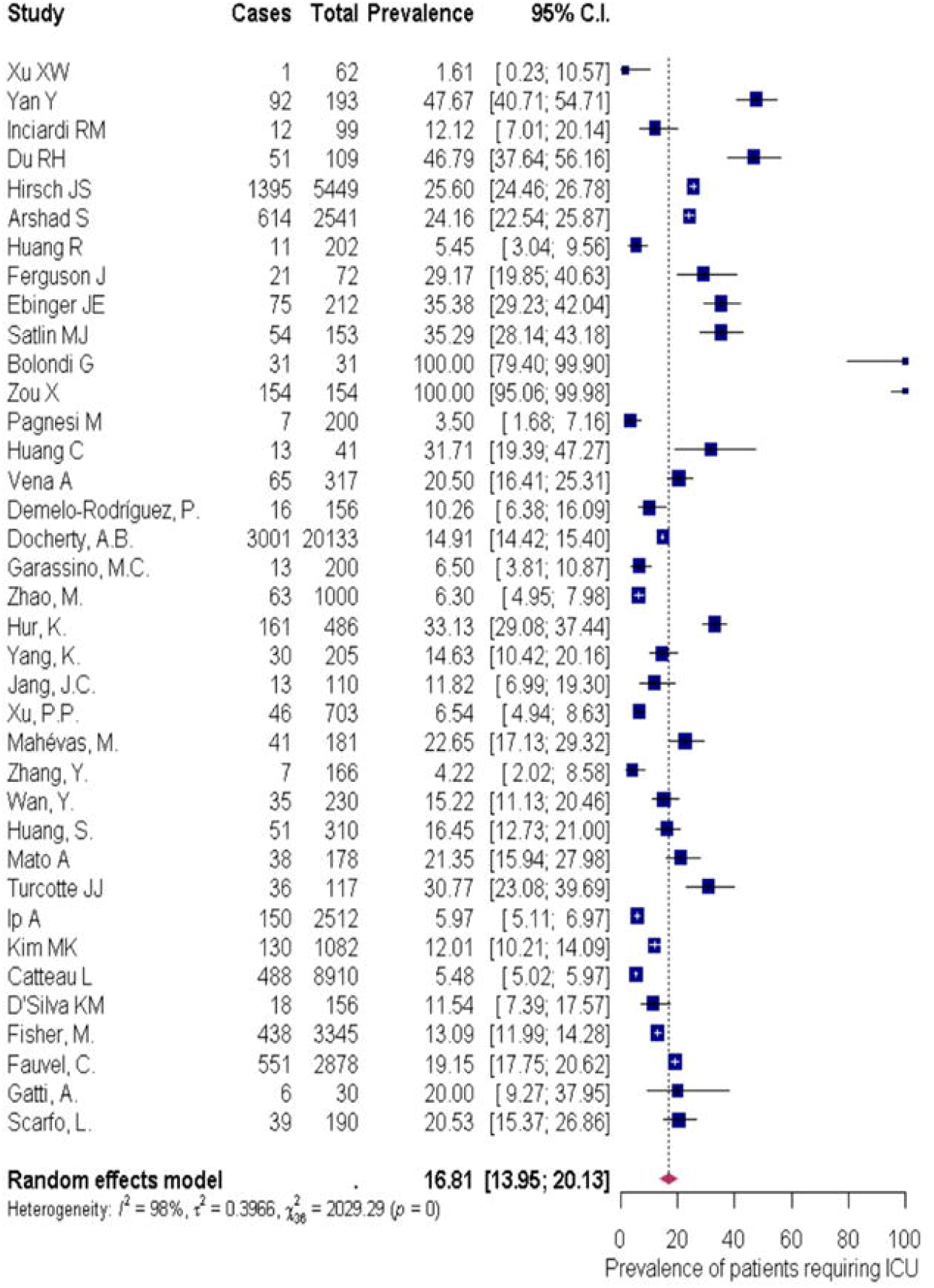
Forest plot of prevalence of admission in the Intensive Care Unit among adult patients with COVID-19 hospitalized in general wards. Only studies with zero and low uncertainty have been included. All included studies have been published in 2020. Abbreviation: CI, confidence interval.

### Secondary endpoints

We described the prevalence of different organ dysfunctions, of admission in the ICU, and of organ support and/or replacement among the 104 studies included. A synthesis of the estimates of prevalence of organ dysfunctions in adults is presented in eTables 3 and 4; similar pediatric data are presented in eTable 5. Acute Respiratory Distress Syndrome (ARDS) was the most common organ dysfunction both in non-ICU and ICU patients. ARDS prevalence in non-ICU wards was 27.6% (95% CI, 21.6-34.5; I^2^, 99%) whereas in ICU reached 88.3% (95% CI, 79.7-93.5; I^2^, 97%; p<0.0001). Shock was the second most common dysfunction in ICU patients, followed by renal and liver dysfunction (Table 1, eFigure 5). Coagulopathy was the second most common organ dysfunction among non-ICU patients. Prevalence of all organ dysfunctions was much higher in ICU than non-ICU patients except for coagulopathy and central nervous system dysfunction; the prevalence of these dysfunctions interestingly remained similar.

**Table 1.** Summary of the pooled estimates of prevalence of organ dysfunctions among adult patients hospitalized in the Intensive Care Unit (ICU) and non-ICU wards.

| Dysfunction | Non-ICU patients |  |  |  |  |  | ICU patients |  |  |  |  |  |
| --- | --- | --- | --- | --- | --- | --- | --- | --- | --- | --- | --- | --- |
|  | N of studies | N of Patients | N with dysfunction | Prevalence, % | 95% CI | I<sup>2</sup>, % | N of studies | N of patients | N with dysfunction | Prevalence, % | 95% CI | I<sup>2</sup>, % |
| **ARDS** | 34 | 26644 | 5555 | 27.6 | 21.6-34.5 | 99 | 25 | 3661 | 2124 | 88.3 | 79.7-93.5 | 97 |
| **Shock** | 23 | 16455 | 1775 | 7.2 | 4.6-11.0 | 96 | 15 | 2752 | 803 | 34.4 | 23.9-46.7 | 96 |
| **Renal dysfunction** | 36 | 31898 | 4700 | 11.7 | 7.7-17.3 | 99 | 19 | 2912 | 673 | 22.0 | 15.0-31.0 | 95 |
| **Coagulopathy** | 34 | 14615 | 1530 | 13.0 | 10.1-16.5 | 95 | 21 | 3398 | 501 | 12.5 | 7.7-19.8 | 95 |
| **Liver dysfunction** | 18 | 10977 | 690 | 6.8 | 4.4-10.3 | 94 | 14 | 2566 | 411 | 18.1 | 9.1-32.7 | 96 |
| **CNS dysfunction** | 13 | 29548 | 4856 | 5.4 | 3.3-8.5 | 98 | 4 | 1520 | 57 | 4.7 | 1.3-16.2 | 96 |

Abbreviations: ARDS acute respiratory distress syndrome; CI confidence interval; CNS central nervous system; ICU intensive care unit; N number.

Among adult patients hospitalized in the general ward, 16.9% (95% CI, 14.0-20.1; I^2^, 96%) required ICU admission (Figure 3). As expected, need for organ replacement was more likely among ICU than non-ICU patients (Table 2). Pooled estimates of patients in the ICU requiring mechanical ventilation (eFigure 6A), renal replacement (eFigure 6B), and ECMO (eFigure 6C) were 57.3% (95% CI, 44.1-69.5; I^2^, 97%), 14.1% (95% CI, 9.5-20.3; I^2^, 91%), and 6.7% (95% CI, 3.5-12.6; I^2^, 93%), respectively.

**Table 2.** Summary of the pooled estimates of prevalence of organ replacement among adult patients hospitalized in the Intensive Care Unit (ICU) and non-ICU wards.

| Type of replacement | Non-ICU patients |  |  |  |  |  | ICU patients |  |  |  |  |  |
| --- | --- | --- | --- | --- | --- | --- | --- | --- | --- | --- | --- | --- |
|  | N of studies | N of patients | N with replacement | Prevalence, % | 95% CI | I<sup>2</sup>, % | N of studies | N of patients | N with replacement | Prevalence, % | 95% CI | I<sup>2</sup>, % |
| **NIMV** | 25 | 41519 | 4185 | 10.6 | 7.7-14.5 | 96 | 20 | 3448 | 849 | 31.3 | 20.4-44.6 | 97 |
| **MV** | 43 | 65084 | 7714 | 11.1 | 9.3-13.3 | 98 | 27 | 4495 | 1881 | 57.3 | 44.1-69.5 | 97 |
| **ECMO** | 18 | 21631 | 191 | 1.1 | 0.7-1.8 | 82 | 17 | 3032 | 144 | 6.7 | 3.5-12.6 | 93 |
| **CRRT/**<br>**dialysis** | 16 | 9048 | 283 | 3.1 | 1.9-5.0 | 90 | 14 | 2706 | 336 | 14.1 | 9.5-20.3 | 91 |

Abbreviations: CI confidence interval; CRRT continuous renal replacement therapy; ECMO extracorporeal membrane oxygenation;
ICU intensive care unit; MV mechanical ventilation; NIMV non-invasive mechanical ventilation; N number.

Mortality could not be assessed separately for patients with and without sepsis. Nevertheless, Yang X. et al, reported that median SOFA of survivors was 4 (interquartile range [IQR], 3-4) compared to 6 (IQR, 4-8) for non-survivors, showing an indirect effect of sepsis severity on final outcome (20). Similar results were reported by others (33, 73). Zou X et al, estimated the hazard ratio (HR) of SOFA score for death 1.40 (95%CI, 1.28-1.54) (37).

Finally, selected literature was searched for the presence of alterations in inflammation and coagulation markers. Reported markers are summarized in eTables 6 and 7. Lymphopenia was reported in most studies, as well as elevation of acute phase proteins such as ferritin and C-reactive protein (CRP). Ferritin was frequently (7 of 14 studies) elevated with cocentrations higher than 1,000ng/mL, whereas the elevation of CRP was more inconsistent. Procalcitonin remained low in almost all cases, as expected in viral infections. The coagulation pathway was also in many studies affected with elevation of fibrinogen and d-dimers, whereas prothrombin time (PT) and activated partial thromboplastin time (aPTT) interestingly hardly ever deviated above the upper normal limit.

## DISCUSSION

In this systematic review and meta-analysis, we showed that sepsis, based on Sepsis-3 criteria, is present in a considerable proportion of hospitalized patients with COVID-19. Sepsis affects 25% of adult COVID-19 patients outside the ICU and up to 83.8% of those admitted in the ICU. In hospitalized children with COVID-19, sepsis is as high as 7.8%. ARDS is the most common organ dysfunction both in ICU and non-ICU patients.

Sepsis in COVID-19 is more frequently implied, rather than reported. Already in the first published reports, COVID-19 was identified as a potential cause of multiple organ failure leading to death. Chen et al, reported confusion in 9%, acute kidney injury in 3%, shock in 4%, and liver dysfunction in about 50% of patients. Interestingly, bacterial or fungal coinfection was present only in 1% and 4% of cases, respectively, suggesting that SARS-CoV-2 was the main cause of infection-related organ dysfunction, although the incidence of opportunistic infections is likely to rise after the widespread introduction of dexamethasone as treatment (4). Similar results were reported by other studies (5). Viral sepsis has also been associated with influenza A (ARDS present in 72.6% and shock in 32.7% of critically ill patients with H1N1 and H3N2), but variably attributed to secondary bacterial infections, which is not the case in SARS-CoV-2 (123,124).

Pediatric data were obviously sparse. Children are less affected by the virus, estimated as 1-5% of total cases and if ill, they present with milder symptoms. Although prevalence of sepsis was lower than in adults, it was still higher than expected based on initial reports (125).

Key mechanisms that may have a role in the pathophysiology of multi-organ injury secondary to infection with SARS-CoV-2 include direct viral toxicity, endothelial cell damage and thrombo-inflammation, dysregulation of the immune response, and dysregulation of the renin–angiotensin–aldosterone system (RAAS) (126). Some of these mechanisms may be unique to COVID-19 such as that multiple-organ injury may occur at least in part due to direct viral tissue damage (127,128). The host response with activation of the adaptive and innate immune system and the resulting systemic release of cytokines and activation of the complement system, are common features of both bacterial and viral sepsis (126,129,130).

Severe COVID-19 has been shown to display a similar cytokine profile in terms of baseline levels of interleukin (IL)-1β, IL-1RA, IL-6, IL-8, IL-18, and TNF-α, as ARDS of other etiology and bacterial sepsis, although the absolute circulating concentrations tend to be somewhat lower (131). The presence of macrophage activation-like syndrome, an extreme phenotype of bacterial sepsis, characterized by hyperferritinemia, coagulation disorders, liver dysfunction and high mortality, is also present in severe COVID-19. (8,132,133). Meanwhile, it has been demonstrated that the host response is characterized by a dysregulated activation of the innate immune system coupled with tissue damage by neutrophils and monocytes driving the interplay of inflammation and coagulation (129,130). Altogether, these observations suggest that death from COVID-19 to significant degree is a consequence of immune-mediated organ damage, rather than a result of the infection itself (3,8).

Following the paradigm of bacterial sepsis and since no specific antiviral treatment is available for COVID-19, immune targeting therapies have been tested through several clinical trials; corticosteroids, IL-1 and IL-6 blockade and inhibition of factors of the complement system have been administered to patients with organ dysfunctions with promising results (134-139). Similarities between sepsis and COVID-19 have been early recognized by sepsis experts, issuing a written guide for appropriate management of COVID-19 (140).

To the best of our knowledge, this is the first study to address in a systematic way the presence of sepsis, according to Sepsis-3 criteria, among hospitalized patients with COVID-19, and the first to provide pooled estimates of specific organ dysfunctions. Of course, the conservative approach used in SOFA score extraction (i.e. median or mean SOFA of included cohorts) may have resulted in an underestimation of sepsis prevalence. Sensitivity analyses focusing on low uncertainty data provided similar prevalence, reducing this potential bias. High heterogeneity was observed in almost all results, which could not be eliminated by sensitivity analyses. This can be explained by the heterogeneity in reporting and definitions of organ dysfunctions in the various studies, a shift in criteria for hospitalization since the beginning of the pandemic (e.g. mild cases tend to receive home care). Above all, it is likely that COVID-19-associated sepsis, similar to bacterial sepsis and ARDS, is a heterogenous disease and every effort should be made to identify patients at high risk for those complications.

In conclusion, a considerable proportion of patients with COVID-19 present viral sepsis. Lessons learned from bacterial sepsis may apply, in terms of early recognition (by means of SOFA and/or qSOFA score) and potential benefit from immune regulating strategies.

## Data Availability

Data are available upon reasonable request. Address correspondence to

## Contributors

E.Ky. performed literature search and study selection, participated in data analysis and drafted the manuscript. E.Ka. performed literature search and study selection and participated in data analysis and drafting of the manuscript. M.K. performed data analysis. C.F.S., M.G.N. and K.R. conceptualized the study and revised the manuscript for important intellectual content. E.J.G.B. conceptualized the study, participated in literature search, study selection and drafting the manuscript. All authors gave approval for the version to be published.

## Acknowledgements

There was no funding source for this study.

The authors would like to thank following researchers/scientists for providing clinical data for this study: Dr. Marco Metra, Dr. Laura Lupi, Dr. Calum Semple, Dr. Annemarie Docherty, Dr. Stephen Knight, Dr. Donald E Griesdale, Dr. Nick Fergusson, Dr. Matteo Pagnesi, Dr. Furkan Korkmaz, Dr. Matthijs Kox, Dr. Giuliano Bolondi, Dr. Margaux Lafaurie and Dr. Guillaume Moulis.

