## Supplement for "COVID-19 as cause of viral sepsis: A Systematic Review and Meta-Analysis"

**“Online Supplement”**

Eleni Karakike, Evangelos J. Giamarellos-Bourboulis, Miltiades Kyprianou, Carolin Fleischmann-Struzek, Mihai Netea, Konrad Reinhart, Evdoxia Kyriazopoulou

**Table of contents**

| **Content** | **Page** |
| --- | --- |
| **Search Strategy.** | 3 |
| **Supplementary Figures.** | 5 |
| **eFigure 1.** Prevalence of sepsis and 95% Confidence Intervals (CI). | 5 |
| **eFigure 2.** Funnel plot of publication bias for trials included in analysis of prevalence of sepsis A) among adults outside the Intensive Care Unit (ICU); B) among adults in the ICU; and C) among pediatric patients outside the ICU. | 6 |
| **eFigure 3.** Sensitivity analysis. Prevalence of sepsis and 95% Confidence Intervals (CI). Only adult studies of which corresponding authors provided clinical data are included; A) outside the Intensive Care Unit (ICU) and B) in the ICU. Corresponding funnel plots of publication bias for studies outside (C) and in the ICU (D) are presented. | 7 |
| **eFigure 4.** Sensitivity analysis. Prevalence of sepsis and 95% Confidence Intervals (CI). Only adult studies of which high uncertainty was assessed are included; A) outside the Intensive Care Unit (ICU) and B) in the ICU. Corresponding funnel plots of publication bias for studies outside (C) and in the ICU (D) are presented. | 8 |
| **eFigure 5.** Prevalence of shock and 95% Confidence Intervals (CI) A) among adult patients treated in general ward and B) among adult patients treated in an Intensive Care Unit (ICU) | 19 |
| **eFigure 6.** Forest plot of prevalence and 95% Confidence Interval (CI) of A) mechanical ventilation; B) renal replacement; C) extracorporeal membrane oxygenation (ECMO) among adult patients with COVID-19 hospitalized in the Intensive Care Unit.1 | 10 |
| **Supplementary Tables.** | 11 |
| **eTable 1**. Characteristics of the included studies. | 11 |
| **eTable 2.** Risk assessment of included studies according to MINORS score. | 16 |
| **eTable 3.** Prevalence of organ dysfunctions in adult patients with COVID-19 hospitalized outside the Intensive Care Unit, as reported in included studies. | 19 |
| **eTable 4.** Prevalence of organ dysfunctions in adult patients with COVID-19 in the Intensive Care Unit, as reported in included studies. | 21 |
| **eTable 5.** Prevalence of specific organ dysfunctions in children hospitalized with COVID-19, as reported in included studies. | 23 |
| **eTable 6.** Summary of inflammation markers in the included studies. | 24 |
| **eTable 7.** Summary of coagulation markers in the included studies. | 26 |

**Search Strategy.**

| **PubMed (Date Run: 03/10/2020)** | | |
| --- | --- | --- |
| Step | **Search strategy** | **Found** |
| **#1** | [(Covid AND sepsis) OR (Covid AND septic*) OR (Covid and "organ failure") OR (Covid and "organ dysfunction") OR (Covid AND "organ support") OR (Covid AND SOFA) OR (Covid and "intensive care") OR (Covid AND "organ replacement") OR (Covid AND "host response") OR (Covid AND "immune response") OR (Covid AND "renal failure") OR (Covid AND "renal dysfunction") OR (Covid AND „pulmonary failure") OR (Covid AND „pulmonary dysfunction") OR (Covid AND „respiratory failure") OR (Covid AND „respiratory dysfunction") OR (Covid AND „liver failure") OR (Covid AND „liver dysfunction") OR (Covid AND „cardiac failure") OR (Covid AND „cardiac dysfunction") OR (Covid AND „shock") OR (Covid AND „neurologic dysfunction") OR (Covid AND „neurological dysfunction") OR (Covid AND thrombocytopenia) OR (Covid AND acidosis)] OR [(SARS-CoV-2 AND sepsis) OR (SARS-CoV-2 AND septic*) OR (SARS-CoV-2 AND "organ failure") OR (Covid and "organ dysfunction") OR (SARS-CoV-2 AND "organ support") OR (SARS-CoV-2 AND SOFA) OR (SARS-CoV-2 AND "intensive care") OR (SARS-CoV-2 AND "organ replacement") OR (SARS-CoV-2 AND "host response") OR (SARS-CoV-2 AND "immune response") OR (SARS-CoV-2 AND "renal failure") OR (SARS-CoV-2 AND "renal dysfunction") OR (SARS-CoV-2 AND „pulmonary failure") OR (SARS-CoV-2 AND „pulmonary dysfunction") OR (SARS-CoV-2 AND „respiratory failure") OR (SARS-CoV-2 AND „respiratory dysfunction") OR (SARS-CoV-2 AND „liver failure") OR (SARS-CoV-2 AND „liver dysfunction") OR (SARS-CoV-2 AND „cardiac failure") OR (SARS-CoV-2 AND „cardiac dysfunction") OR (SARS-CoV-2 AND „shock") OR (SARS-CoV-2 AND „neurologic dysfunction") OR (SARS-CoV-2 AND „neurological dysfunction") OR (SARS-CoV-2 AND thrombocytopenia) OR (SARS-CoV-2 AND acidosis)] Filters: Full text, Clinical Study, Clinical Trial, Clinical Trial, Phase I, Clinical Trial, Phase II, Clinical Trial, Phase III, Clinical Trial, Phase IV, Comparative Study, Controlled Clinical Trial, Multicenter Study, Observational Study, Pragmatic Clinical Trial, Randomized Controlled Trial, Humans, English | 325 |

| **Cochrane Central Register of Clinical Trials (Date Run: 03/10/20)** | | |
| --- | --- | --- |
| ID | **Search strategy** | **Found** |
| **#1** | (COVID-19):ti,ab,kw OR ("SARS Co-V"):ti,ab,kw | 1571 |
| **#2** | ("sepsis"):ti,ab,kw OR ("organ damage"):ti,ab,kw OR ("organ failure"):ti,ab,kw OR ("sofa"):ti,ab,kw OR ("intensive care"):ti,ab,kw | 33832 |
| **#3** | ("immune response"):ti,ab,kw OR ("host reaction"):ti,ab,kw | 8913 |
| **#4** | ("renal failure"):ti,ab,kw OR ("renal dysfunction"):ti,ab,kw | 8567 |
| **#5** | ("respiratory failure"):ti,ab,kw OR ("respiratory distress"):ti,ab,kw | 10001 |
| **#6** | ("liver damage"):ti,ab,kw OR (liver replacement):ti,ab,kw | 1277 |
| **#7** | ("cardiac failure"):ti,ab,kw OR ("cardiovascular collapse"):ti,ab,kw | 942 |
| **#8** | ("shock"):ti,ab,kw OR ("acidosis"):ti,ab,kw | 12526 |
| **#9** | ("neurological impairment"):ti,ab,kw OR ("encephalopathy"):ti,ab,kw | 3530 |
| **#10** | ("thrombocytopenia"):ti,ab,kw OR ("thrombopenia"):ti,ab,kw | 9554 |
| **#11** | #2 OR #3 OR #4 OR #5 OR #6 OR #7 OR #8 OR #9 OR #10 | 76561 |
| **#12** | #1 AND #11 | 501 |

| **Google Scholar (Date Run: 03/10/20)** | | |
| --- | --- | --- |
| ID | **Search strategy** | **Found** |
| **#1** | [(Covid AND sepsis) OR (Covid AND septic*) OR (Covid and "organ failure") OR (Covid and "organ dysfunction") OR (Covid AND "organ support") OR (Covid AND SOFA) OR (Covid and "intensive care") OR (Covid AND "organ replacement") OR (Covid AND "host response") OR (Covid AND "immune response") OR (Covid AND "renal failure") OR (Covid AND "renal dysfunction") OR (Covid AND „pulmonary failure") OR (Covid AND „pulmonary dysfunction") OR (Covid AND „respiratory failure") OR (Covid AND „respiratory dysfunction") OR (Covid AND „liver failure") OR (Covid AND „liver dysfunction") OR (Covid AND „cardiac failure") OR (Covid AND „cardiac dysfunction") OR (Covid AND „shock") OR (Covid AND „neurologic dysfunction") OR (Covid AND „neurological dysfunction") OR (Covid AND thrombocytopenia) OR (Covid AND acidosis)] OR [(SARS-CoV-2 AND sepsis) OR (SARS-CoV-2 AND septic*) OR (SARS-CoV-2 AND "organ failure") OR (Covid and "organ dysfunction") OR (SARS-CoV-2 AND "organ support") OR (SARS-CoV-2 AND SOFA) OR (SARS-CoV-2 AND "intensive care") OR (SARS-CoV-2 AND "organ replacement") OR (SARS-CoV-2 AND "host response") OR (SARS-CoV-2 AND "immune response") OR (SARS-CoV-2 AND "renal failure") OR (SARS-CoV-2 AND "renal dysfunction") OR (SARS-CoV-2 AND „pulmonary failure") OR (SARS-CoV-2 AND „pulmonary dysfunction") OR (SARS-CoV-2 AND „respiratory failure") OR (SARS-CoV-2 AND „respiratory dysfunction") OR (SARS-CoV-2 AND „liver failure") OR (SARS-CoV-2 AND „liver dysfunction") OR (SARS-CoV-2 AND „cardiac failure") OR (SARS-CoV-2 AND „cardiac dysfunction") OR (SARS-CoV-2 AND „shock") OR (SARS-CoV-2 AND „neurologic dysfunction") OR (SARS-CoV-2 AND „neurological dysfunction") OR (SARS-CoV-2 AND thrombocytopenia) OR (SARS-CoV-2 AND acidosis) | 1077 |

**Supplementary Figures.**

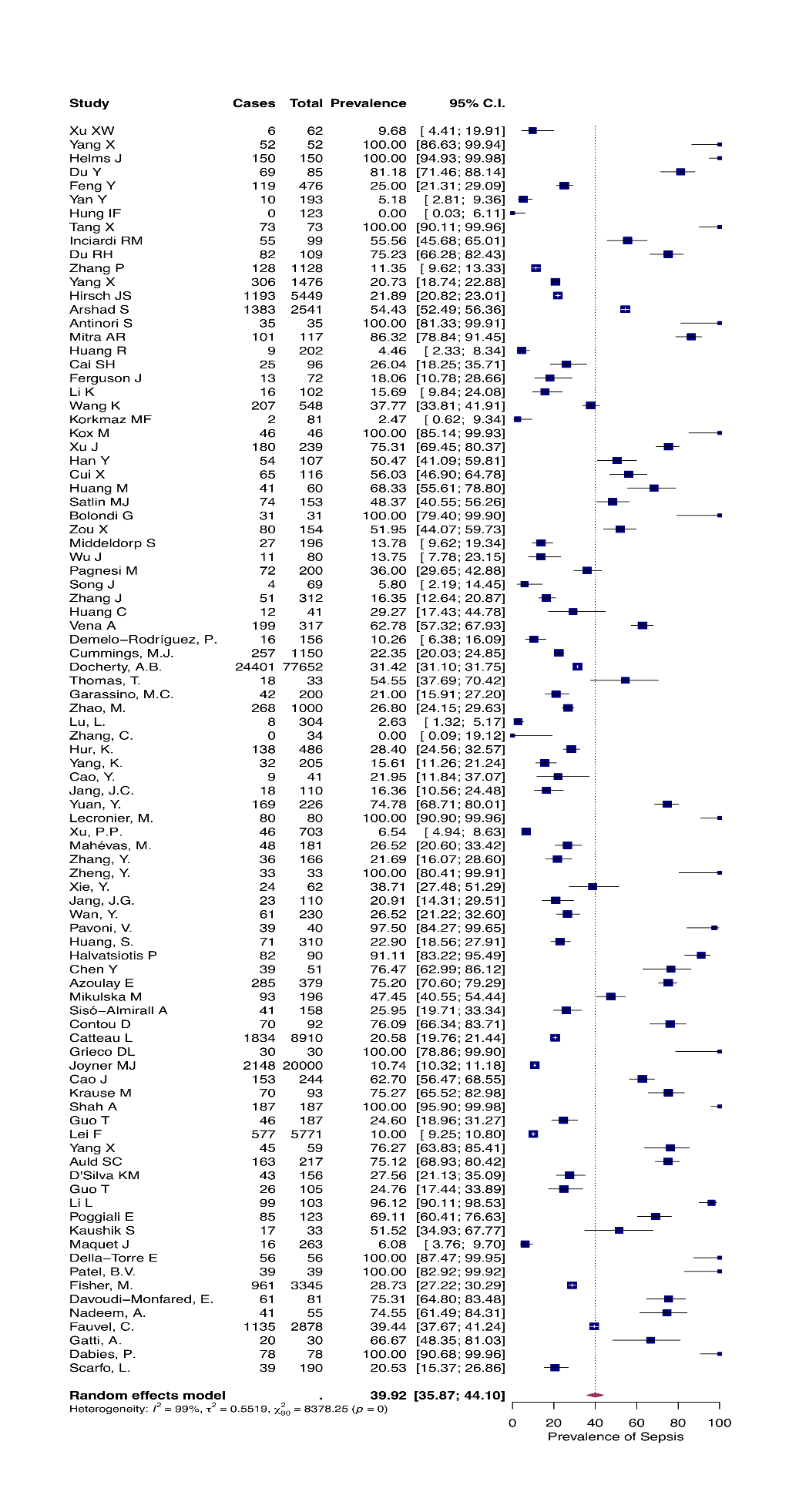
**eFigure 1.** Prevalence of sepsis and 95% Confidence Intervals (CI). All 92 articles providing available data were included in analysis.

**eFigure 2.** Funnel plot of publication bias for trials included in the primary analysis on sepsis prevalence A) among adults outside the Intensive Care Unit (ICU); B) among adults in the ICU; and C) among pediatric patients outside the ICU.

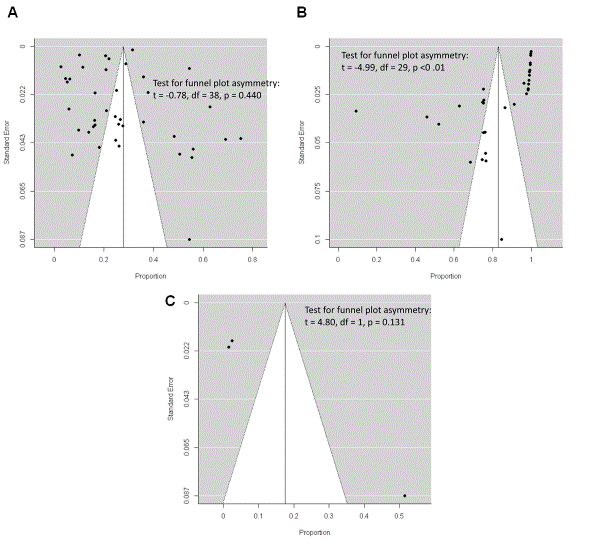

**eFigure 3.** Sensitivity analysis. Prevalence of sepsis and 95% Confidence Intervals (CI). Only adult studies of zero uncertainty are included; A) outside the Intensive Care Unit (ICU) and B) in the ICU. Corresponding funnel plots of publication bias for studies outside (C) and in the ICU (D) are presented.

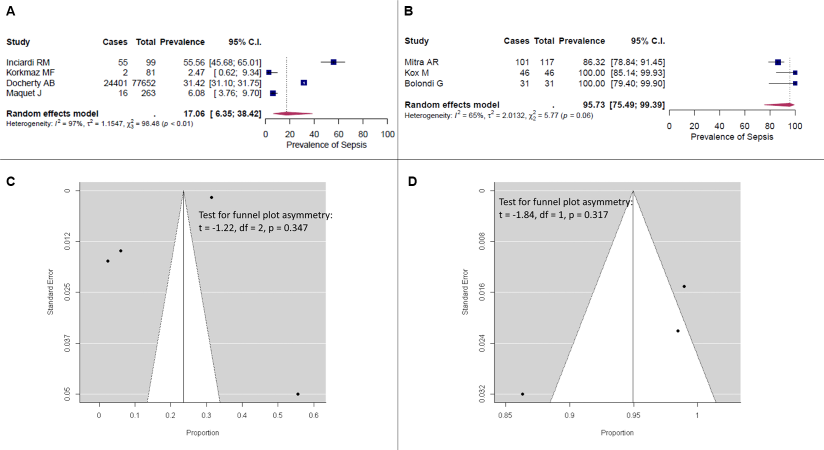

**eFigure 4.** Sensitivity analysis. Prevalence of sepsis and 95% Confidence Intervals (CI). Only adult studies of high uncertainty are included; A) outside the Intensive Care Unit (ICU) and B) in the ICU. Corresponding funnel plots of publication bias for studies outside (C) and in the ICU (D) are presented.

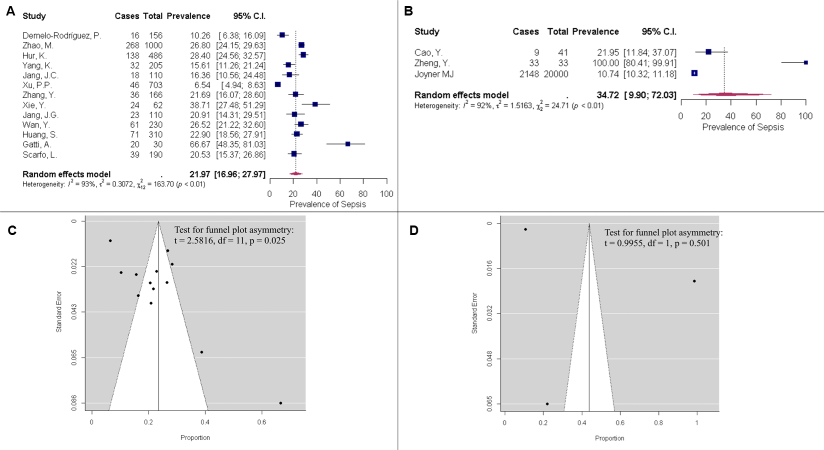

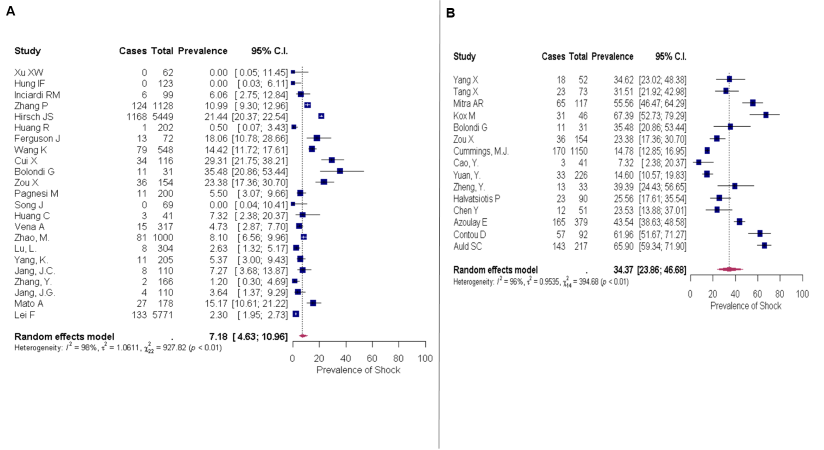
**eFigure 5.** Prevalence of shock and 95% Confidence Intervals (CI) A) among adult patients treated in general ward and B) among adult patients treated in an Intensive Care Unit (ICU).

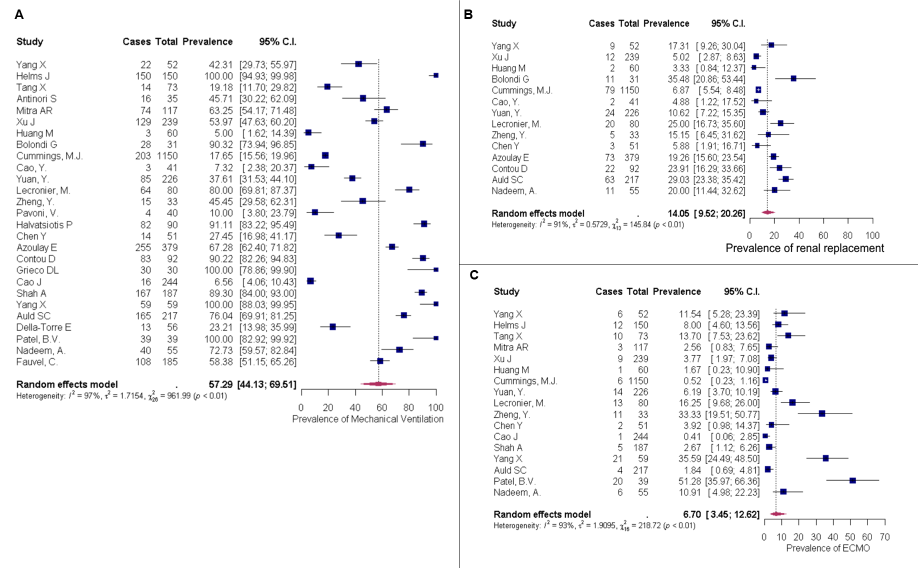
**eFigure 6.** Forest plot of prevalence and 95% Confidence Interval (CI) of A) mechanical ventilation; B) renal replacement; C) extracorporeal membrane oxygenation (ECMO) among adult patients with COVID-19 hospitalized in the Intensive Care Unit.

**eTable 1**. Characteristics of the included studies.

| **Ref.** | **Clearly stated aim** | **Publication time** | **Study design** | **Study setting** | **N of patients** | **Incidence of sepsis %** | **Inclusion of consecutive patients** | **Level of uncertainty** |
| --- | --- | --- | --- | --- | --- | --- | --- | --- |
| 19 | Xu XW. | Feb 20 | Retrosp. observational | single-center, Dec 19-Jan 20, China (Zhejiang province) | 62 | 9.7 | Yes – all comers | ** |
| 20 | Yang X. | May 20 | Retrosp. observational | single-center, Dec 19-Jan 20, China (Wuhan) | 52 | 100.0 | Critically ill | ** |
| 109 | Helms J. | Jun 20 | Retrosp. observational | multicenter, Mar 20, France (Strasbourg) | 150 | 100.0 | ARDS | ** |
| 21 | Du Y. | Jun 20 | Retrosp. observational | two-center, Jan 20-Feb 20, China (Wuhan) | 85 | 81.2 | Non-survivors | *** |
| 22 | Dai M. | Jun 20 | Retrosp. observational | multicenter, Jan 20-Feb 20, China (Wuhan) | 641 | NA | Cancer patients | **** |
| 23 | Feng Y. | Jun 20 | Retrosp. observational | multicenter, Jan 20-Feb 20, China (Shanghai, Anhui, Wuhan) | 476 | 25.0 | Yes – all comers | *** |
| 24 | Yan Y. | Apr 20 | Retrosp. observational | single-center, Jan 20-Feb 20, China (Wuhan) | 193 | 5.2 | Yes – all comers | *** |
| 110 | Hung IF. | May 20 | Open-label, phase 2 RCT lopinavir/  ritonavir/ribavirin | multicenter, Feb 20-Mar 20, Hong Kong | 123 | 0.0 | NEWS_2_ score>1 | ** |
| 25 | Tang X. | Jul 20 | Retrosp. case control (comparison with H1N1) | Dec 19-Feb 20, China (Wuhan) | 73 | 100.0 | ARDS | ** |
| 85 | Inciardi RM. | May 20 | Retrosp. observational | single-center, Mar 20, Italy (Brescia, Lombardy) | 99 | 55.6 | Yes – all comers | * |
| 26 | Du RH. | Jul 20 | Observational | multicenter, Dec 19-Feb 20, China (Wuhan) | 109 | 75.2 | Decedents | ** |
| 27 | Zhang P. | Jun 20 | Retrosp. observational | multicenter, Dec 20-Feb 20, China (Hubei) | 1128 | 11.3 | Hypertension | *** |
| 28 | Yang X. | Jun 20 | Retrosp. observational | single-center, Dec 20-Feb 20, China (Wuhan) | 1476 | 20.7 | Yes – all comers | *** |
| 61 | Hirsch JS. | Jul 20 | Retrosp. observational | multicenter, Mar 20-Apr 20, USA | 5449 | 21.9 | Yes – all comers | *** |
| 62 | Arshad S. | Aug 20 | Comparative retrospective for HCQ, AZM or combination | multicenter, Mar 20-May 20, USA (Michigan) | 2541 | 54.4 | Yes – all comers | ** |
| 86 | Antinori S. | Aug 20 | Open-label prospective trial of remdesivir | single-center, Feb 20-Mar 20, Italy (Milan) | 35 | 100.0 | On MV | *** |
| 111 | Mitra AR. | May 20 | Retrosp. case series | Feb 20-Apr 20, Canada (Vancouver) | 117 | 86.3 | Yes – all comers | * |
| 29 | Huang R. | May 20 | Retrosp. observational | multicenter, Jan 20-Feb 20, China (Jiangsu province) | 202 | 4.5 | Yes – all comers | *** |
| 30 | Cai SH. | Jun 20 | Retrosp. observational | multicenter, Jan 20-Feb 20, China | 96 | 26.0 | Yes – all comers | *** |
| 63 | Ferguson J. | Aug 20 | Retrosp. observational | two-center, Mar 20-Apr 20, USA (California) | 72 | 18.1 | Yes – all comers | *** |
| 31 | Li K. | Jun 20 | Retrosp. observational | single-center, Jan 20-Mar 20, China (Wuhan) | 102 | 15.7 | Yes – all comers | *** |
| 32 | Wang K. | Apr 20 | Retrosp. observational | single-center,China | 548 | 37.8 | Yes – all comers | *** |
| 87 | Kox M. | Sep 20 | Retrosp. observational | single-center, Mar 20-Apr 20, Netherlands | 46 | 100.0 | ARDS | * |
| 33 | Xu J. | Jul 20 | Retrosp. observational | multicenter, Jan 20-Feb 20, China (Wuhan) | 239 | 75.3 | Critically ill | ** |
| 34 | Han Y. | Jun 20 | Retrosp. observational | single-center, Feb 20-Mar 20, China (Wuhan) | 107 | 50.5 | Yes – all comers | ** |
| 64 | Ebinger JE. | Jul 20 | Retrosp. observational | multicenter, Feb 20-Mar 20, USA (California) | 212 | NA | Yes – all comers | **** |
| 35 | Cui X. | Jun 20 | Retrosp. observational | multicenter, Jan20-Mar20, China (Wuhan) | 116 | 56.0 | Yes – all comers | ** |
| 36 | Huang M. | Aug 20 | Retrosp. observational | multicenter, Jan20-Apr20, China (Jiangsu province) | 60 | 68.3 | Severe illness | *** |
| 65 | Satlin MJ. | Jul 20 | Retrosp. observational | Mar 20, USA (Manhattan) | 153 | 48.4 | Treatment with HCQ | *** |
| 88 | Bolondi G. | Jun 20 | Retrosp. observational | single-center, Mar20-Apr20, Italy (Cesegna) | 31 | 100.0 | Yes – all comers | ** |
| 37 | Zou X. | Aug 20 | Retrosp. observational | single-center, Jan20-Feb20, China (Wuhan) | 154 | 51.9 | Yes – all comers | ** |
| 89 | Middeldorp S. | Aug 20 | Retrosp. observational | single-center, Apr 20, Netherlands | 123 | 16.3 | Yes – all comers | *** |
| 38 | Wu J. | Jul 20 | Retrosp. observational | multicenter, Jan20-Feb20, China (Jiangsu Province) | 80 | 13.8 | Yes – all comers | *** |
| 90 | Pagnesi M. | Sep 20 | Cross-sectional | single-center, Mar 20-Apr 20, Italy (Milan) | 200 | 36.0 | Yes – all comers | *** |
| 39 | Song J. | Jul 20 | Retrosp. observational | single-center, Jan 20-Feb20, China (Hubei) | 69 | 5.8 | Yes – all comers | *** |
| 40 | Zhang J. | Jul 20 | Retrosp. observational | multicenter, Jan 20-Mar 20, China (Wuhan) | 312 | 16.3 | Yes – all comers | *** |
| 41 | Huang C. | Jan 20 | Retrosp. observational | single-center, Dec 20-Jan 20, China (Wuhan) | 28 | 7.1 | Yes – all comers | *** |
| 91 | Vena A. | Aug 20 | Retrosp. observational | single-center, Feb 20-Mar 20, Italy (Genoa) | 317 | 62.8 | Yes – all comers | *** |
| 92 | Demelo-Rodríguez P. | Aug 20 | Prosp. observational | single-center, Apr 20, Spain (Madrid) | 156 | 10.3 | High D-dimers | **** |
| 66 | Cummings MJ. | Jun 20 | Prosp. observational | two-center, Mar 20-April 20, USA (New York) | 1,152 | 22.3 | Critical illness | ** |
| 80 | Docherty AB. | Μay 20 | Prosp. observational | multi-center, Feb 20-Apr 20  UK (Wales, England, Scotland) | 20133 | 21.5 | Yes – all comers | * |
| 67 | Thomas T. | Jul 20 | Observational | Not reported  USA (New York) | 33 | 54.5 | Yes – all comers | *** |
| 120 | Garassino MC. | Jul 20 | Observational | multicenter, Mar 20-Apr 20  Italy, Spain, France, Switzerland, Netherlands, USA, UK and China | 200 | 21.0 | Cancer patients | *** |
| 42 | Zhao M. | Jun 20 | Retrosp. observational | single-center, Jan 20-Feb 20, China (Wuhan) | 1000 | 26.8 | Yes – all comers | **** |
| 43 | Lu L. | Jun 20 | Retrosp. observational | multicenter, Jan 20-Feb 20, China (Sichuan, Hubei province, Chongqing mun) | 304 | 2.6 | Yes – all comers | *** |
| 68 | Hur K. | Jul 20 |  | multicenter, Mar 20-Apr 20, USA (Chicago) | 486 | 28.4 | Yes – all comers | **** |
| 112 | Yang K. | Jul 20 | Retrosp. observational | Multicenter, Jan 20- Mar 20 | 205 | 15.6 | Cancer patients | **** |
| 44 | Cao Y. | Jul 20 | Single-blind RCT of ruxolitinib | multicenter, Feb 20, China (Hubei, Hunan province) | 41 | 22.0 | Severe illness | **** |
| 113 | Jang JC. | Jun 20 | Retrosp. observational | single-center, Feb 20-Mar 20, South Corea (Daegu) | 110 | 16.4 | Yes – all comers | *** |
| 45 | Yuan Y. | Μay 20 | Cross-sectional | multicenter, Feb 20, China (Wuhan) | 226 | 74.8 | Severe illness | ** |
| 83 | Lecronier M. | Jul 20 | Retrosp. observational | single-center, Mar 20-Apr 20, France (Paris) | 80 | 100.0 | Acute respir. Failure | ** |
| 46 | Xu PP. | Μay 20 | Retrosp. observational | multicenter, Jan 20-Mar 20, China (Hubei, Zhejiang, Shandong, Jiangsu provice) | 703 | 6.5 | Yes – all comers | **** |
| 94 | Mahévas M. | Μay 20 | Retrosp. observational | multicenter, Mar 20, France (Ile-de-France) | 181 | 26.5 | Requiring O_2_ | *** |
| 47 | Zhang Y. | Μay 20 | Retrosp. observational | single-center, Feb 20-Mar 20, China (Wuhan) | 166 | 21.7 | Hyperglycemia | **** |
| 48 | Zheng Y. | Μay 20 | Retrosp. observational | single-center, Jan 20-Mar 20, China (Hangzhou) | 33 | 100.0 | Severe illness | **** |
| 49 | Xie Y. | Aug 20 | Retrosp. observational | single-center, Feb 20-Mar 20, China (Wuhan) | 62 | 38.7 | Yes – all comers | **** |
| 114 | Jang JG. | Jun 20 | Retrosp. observational | single-center, Feb 20-Apr 20, Korea (Daegu) | 110 | 20.9 | Yes – all comers | **** |
| 50 | Wan Y. | Jun 20 | Retrosp. observational | multicenter, Jan 20-Mar 20. China (Guangdong, Hubei, Jiangxi province) | 230 | 26.5 | Yes – all comers | **** |
| 95 | Pavoni V. | Μay 20 | Retrosp. observational | single-center, Feb 20-Apr 20, Italy (Tuscany) | 40 | 97.5 | Severe illness | ** |
| 51 | Huang S. | Jun 20 | Prosp. observational | two-center, China (Wuhan) | 310 | 22.9 | Hypertension | **** |
| 96 | Halvatsiotis P. | Jul 2020 | Retrosp. observational | multicenter, Mar 20-Apr 20, Greece | 90 | 91.1 | ICU | *** |
| 121 | Mato A. | Sep 20 | Retrosp. observational | multicenter, Feb 20-Apr 20, USA, European Union/United Kingdom and South America | 178 | NA | CLL | **** |
| 52 | Chen Y. | Jul 20 | Retrosp. observational | multicenter, Jan 20-Mar 20, China (Hebei) | 51 | 76.5 | Critically ill | ** |
| 97 | Karagiannidis C. | Jul 20 | Retrosp. observational | multicenter, Feb 20-Apr 20, Germany | 10,021 | NA | Yes – all comers | **** |
| 98 | Azoulay E. | Sep 20 | Retrosp. observational | multicenter, Feb 20-Apr 20, France (Paris) | 379 | 75.2 | ICU | ** |
| 69 | Turcotte JJ. | Aug 20 | Retrosp. observational | single center, Mar 20 -Apr 20, USA (Annapolis) | 117 | NA | Yes – all comers | **** |
| 70 | Ip A. | Aug 20 | Retrosp. observational | Multicenter, Mar 20-May 20, USA | 2512 | NA | Yes – all comers | **** |
| 115 | Kim MK. | Aug 20 | Retrosp. observational | Feb 20-Mar 20, South Korea | 1082 | NA | Yes – all comers | **** |
| 99 | Mikulska M. | Aug 20 | Observational trial of tocilizumab/steroids | single-center, Genova, Italy | 196 | 47.4 | Severe illness | *** |
| 100 | Sisó-Almirall A. | Aug 20 | Observational | multicenter, Feb 20-Apr 20 Barcelona, Spain | 158 | 25.9 | Yes – all comers | *** |
| 101 | Contou D. | Aug 20 | Retrosp. observational | single-center, Mar 20-Apr 20, France (Argenteuil) | 92 | 76.1 | Critically ill | ** |
| 102 | Catteau L. | Oct 20 | Retrosp. observational | multicenter, Mar 20-May 20, Belgium | 8910 | 20.6 | Yes – all comers | *** |
| 103 | Grieco DL. | Aug 20 | Prosp, observational | single-center, Mar 20, Italy (Rome) | 30 | 100.0 | ARDS | *** |
| 71 | Joyner MJ. | Sep 20 | Pragmatic trial of convalescent plasma | multicenter, Apr 20-Jun 20,USA | 20,000 | 10.7 | Severe or critical illness | **** |
| 53 | Cao J. | Jul 20 | Retrosp. observational | single-center, Feb 20, China (Wuhan) | 244 | 62.7 | Yes – all comers | *** |
| 72 | Krause M. | Sep 20 | Retrosp. observational | multicenter, Mar 20-Apr 20, USA | 93 | 75.3 | On MV | ** |
| 81 | Shah A. | Sep 20 | Retrosp. observational | multicenter, Mar 20-May 20, UK | 187 | 100.0 | ICU | *** |
| 54 | Guo T. | Jul 20 | Retrosp. observational | single-center, Jan 20-Feb 20, China (Wuhan) | 187 | 24.6 | Yes – all comers | *** |
| 55 | Lei F. | Aug 20 | Retrosp. observational | multicenter, Dec 19-Mar 20, China (Wuhan) | 5771 | 10.0 | Yes – all comers | *** |
| 56 | Yang X. | Sep 20 | Descriptive study | two-center,Jan 20-Mar 20, China (Wuhan) | 59 | 76.3 | On MV | ** |
| 73 | Auld SC. | Sep 20 | Cohort study | multicenter, Mar 20-Apr 20, USA (Atlanda) | 217 | 75.1 | ICU | ** |
| 74 | D'Silva KM. | Sep 20 | Observational | multicenter, Jan 20-Apr 20, USA (Boston, Massachusetts) | 156 | 27.6 | Rheumatologic patients | *** |
| 75 | Maatman TK. | Sep 20 | Observational | multicenter, Mar 20, USA (Indianapolis) | 109 | NA | Yes – all comers | *** |
| 57 | Guo T. | May 20 | Retrosp. observational | multicenter, Jan 20-Feb 20, China (Hunan Province) | 105 | 24.8 | Elderly | *** |
| 58 | Li L. | Jun 20 | Open-label RCT of convalescent plasma | multicenter, Feb 20 -Apr 20, China (Wuhan) | 103 | 96.1 | Severe/critical illness | *** |
| 76 | Imam Z. | Oct 20 | Retrosp. observational | multicenter, Mar 20-Apr 20, USA (Michigan) | 1305 | NA | Yes – all comers | *** |
| 104 | Poggiali E. | Oct 20 | Retrosp. observational | single-center, Feb 20-Mar 20, Italy (Emilia-Romagna) | 123 | 69.1 | Yes – all comers | *** |
| 105 | Maquet J. | Sep 20 | Retrosp. observational | single-center, Mar 20-Apr 20, France (Toulouse) | 263 | NA | Yes – all comers | * |
| 106 | Della-Torre E. | Oct 20 | Open-label observational sarilumab vs control | single-center, Mar 20-Apr 20, Italy (Milan) | 56 | 100.0 | Severe illness | *** |
| 82 | Patel BV. | Sep 20 | Retrosp. observational | single-center, Mar 20-Apr 20, UK (London) | 39 | 100.0 | On MV | ** |
| 77 | Fisher M. | Sep 20 | Retrosp. observational | single-center, Mar 20-Apr 20, USA (New York, Bronx) | 3345 | 28.7 | AKI | *** |
| 116 | Davoudi-Monfared E. | Αug 20 | Open-label RCT  IFN b-1a vs placebo | single-center, Feb 20-Apr 20, Iran (Tehran) | 81 | 75.3 | Severe illness | *** |
| 117 | Nadeem A. | Αug 20 | Retrosp. observational | single-center, Mar 20-May 20, UAE (Abu Dabi) | 55 | 74.5 | Critical illness | ** |
| 107 | Fauvel C. | Jul 20 | Retrosp. observational | multicenter, Feb 20-Apr20, France | 1,240 | 36.0 | CTPA referral | *** |
| 108 | Gatti A. | Αug 20 | Prosp. observational | single-center, Mar 20-May 20, Italy (Milan) | 30 | 66.7 | Yes – all comers | *** |
| 122 | Scarfo L. | Sep 20 | Retrosp. observational | mutli-center, global survey (40 countries), Mar 20 and May 20 | 190 | 20.5 | CLL | **** |
|  | *Pediatric studies* | |  |  |  |  |  |  |
| 78 | Feldstein LR. | Jul 20 | Prospective surveillance | multicenter, Mar 20-May 20, USA | 73 | NA | MIS | **** |
| 118 | Korkmaz MF. | Jun 20 | Retrosp. observational | single-center, Mar 20-May 20, Turkey | 81 | 2.5 | Yes – all comers | * |
| 59 | Li H. | Apr 20 | Retrosp. observational | single-center, Jan 20-Feb 20, China (Wuhan) | 40 | NA | Yes – all comers | **** |
| 60 | Zhang C. | Jun 20 | Retrosp. observational | multicenter, Jan 20-Feb 20, West China | 34 | NA | Mild and moderate illness | **** |
| 119 | Kanburoglu MK. | Oct 20 | Prosp. observational | multicenter, Mar 20-Jun 20, Turkey | 37 | NA | ICU | **** |
| 83 | Swann OV. | Aug 20 | Retrosp. observational | multicenter, Jan 20-Jul 20, UK (Wales, England, Scotland) | 651 | NA | Yes – all comers | **** |
| 79 | Kaushik S. | Sep 20 | Retrosp. observational | single-center, Apr 20-May 20  USA (New York) | 33 | 51.5 | MIS | *** |
| 84 | Dabies P. | Sep 20 | Observational | multicenter, Apr 20-May 20, UK | 78 | 100.0 | PICU | ** |

Abbreviations: AKI Acute Kidney Injury; ARDS Acute Respiratory Distress Syndrome; AZM Azithromycin; CLL Chronic Lymphocytic Leukemia; CTPA Computed Tomography-Pulmonary Angiogram; HCQ Hyrdoxychloroquine; ICU Intensive Care Unit; IFN Interferon; MIS Multisystem Inflammatory Syndrome; MV Mechanical Ventilation; NA non applicable; NEWS National Early Warning Score; PICU Pediatric Intensive Care Unit; RCT Randomized Clinical Trial.

Level of Uncertainty: * zero – contacted authors provided the exact proportion of patients with sepsis-3 based on Sequential Organ Failure Assessment (SOFA) score; ** low – SOFA score was provided in articles as median with respective interquartile range; ***intermediate – proportion of patients with sepsis was extracted by the highest proportion of patients with a specific organ dysfunction raising SOFA score ≥ 2 points; ****high – proportion of patients with sepsis was extracted by the proportion of patients characterized severe or critically ill, or requiring ICU.

**eTable 2.** Risk assessment of included studies according to MINORS score.

| **Ref.** | **Clearly stated aim** | **Inclusion of consecutive patients** | **Prospective data collection** | **Endpoints appropriate to study aim** | **Unbiased assessment of study endpoint** | **Follow-up period appropriate to study aim** | **<5% lost to follow-up** | **Prospective calculation of study size** | **Total**  **(out of 16)** |
| --- | --- | --- | --- | --- | --- | --- | --- | --- | --- |
| 19 | 2 | 2 | 0 | 2 | 2 | 2 | 2 | NA | 12 |
| 20 | 2 | 0 | 0 | 2 | 2 | 2 | 2 | NA | 10 |
| 109 | 2 | 0 | 0 | 2 | 2 | 2 | 2 | NA | 10 |
| 21 | 2 | 0 | 0 | 1 | 1 | 2 | 2 | NA | 8 |
| 22 | 2 | 1 | 0 | 0 | 0 | 2 | 2 | NA | 7 |
| 23 | 2 | 2 | 0 | 1 | 1 | 2 | 2 | NA | 10 |
| 24 | 2 | 2 | 0 | 1 | 1 | 2 | 2 | NA | 10 |
| 110 | 2 | 0 | 2 | 2 | 2 | 2 | 2 | NA | 12 |
| 25 | 2 | 0 | 0 | 2 | 2 | 2 | 2 | NA | 10 |
| 85 | 2 | 2 | 0 | 2 | 2 | 2 | 2 | NA | 12 |
| 26 | 2 | 1 | 0 | 2 | 2 | 2 | 2 | NA | 11 |
| 27 | 2 | 1 | 0 | 1 | 1 | 2 | 2 | NA | 9 |
| 28 | 2 | 2 | 0 | 1 | 1 | 2 | 2 | NA | 10 |
| 61 | 2 | 2 | 0 | 1 | 1 | 2 | 2 | NA | 10 |
| 62 | 2 | 2 | 2 | 2 | 2 | 2 | 2 | NA | 14 |
| 86 | 2 | 0 | 2 | 1 | 1 | 2 | 2 | NA | 10 |
| 111 | 2 | 2 | 0 | 2 | 2 | 2 | 2 | NA | 12 |
| 29 | 2 | 2 | 0 | 1 | 1 | 2 | 2 | NA | 10 |
| 30 | 2 | 2 | 0 | 1 | 1 | 2 | 2 | NA | 10 |
| 63 | 2 | 2 | 0 | 1 | 1 | 2 | 2 | NA | 10 |
| 31 | 2 | 2 | 0 | 1 | 1 | 2 | 2 | NA | 10 |
| 32 | 2 | 2 | 0 | 1 | 1 | 2 | 2 | NA | 10 |
| 87 | 2 | 0 | 0 | 2 | 2 | 2 | 2 | NA | 10 |
| 33 | 2 | 0 | 0 | 2 | 2 | 2 | 2 | NA | 10 |
| 34 | 2 | 2 | 0 | 2 | 2 | 2 | 2 | NA | 12 |
| 64 | 2 | 2 | 0 | 0 | 0 | 2 | 2 | NA | 8 |
| 35 | 2 | 2 | 0 | 2 | 2 | 2 | 2 | NA | 12 |
| 36 | 2 | 0 | 0 | 1 | 1 | 2 | 2 | NA | 8 |
| 65 | 2 | 1 | 0 | 1 | 1 | 2 | 2 | NA | 9 |
| 88 | 2 | 2 | 0 | 2 | 2 | 2 | 2 | NA | 12 |
| 37 | 2 | 2 | 0 | 2 | 2 | 2 | 2 | NA | 12 |
| 89 | 2 | 2 | 0 | 1 | 1 | 2 | 2 | NA | 10 |
| 38 | 2 | 2 | 0 | 1 | 1 | 2 | 2 | NA | 10 |
| 90 | 2 | 2 | 0 | 1 | 1 | 2 | 2 | NA | 10 |
| 39 | 2 | 2 | 0 | 1 | 1 | 2 | 2 | NA | 10 |
| 40 | 2 | 2 | 0 | 1 | 1 | 2 | 2 | NA | 10 |
| 41 | 2 | 2 | 0 | 1 | 1 | 2 | 2 | NA | 10 |
| 91 | 2 | 2 | 0 | 1 | 1 | 2 | 2 | NA | 10 |
| 92 | 2 | 1 | 2 | 0 | 0 | 2 | 2 | NA | 9 |
| 66 | 2 | 0 | 2 | 2 | 2 | 2 | 2 | NA | 12 |
| 80 | 2 | 2 | 2 | 2 | 2 | 2 | 0 | NA | 12 |
| 67 | 2 | 2 | 1 | 1 | 1 | 2 | 2 | NA | 11 |
| 120 | 2 | 1 | 1 | 1 | 1 | 2 | 2 | NA | 10 |
| 42 | 2 | 2 | 0 | 0 | 0 | 2 | 2 | NA | 8 |
| 43 | 2 | 2 | 0 | 1 | 1 | 2 | 2 | NA | 10 |
| 68 | 2 | 2 | 0 | 0 | 0 | 2 | 0 | NA | 6 |
| 112 | 2 | 1 | 0 | 0 | 0 | 2 | 2 | NA | 7 |
| 44 | 2 | 0 | 2 | 0 | 0 | 2 | 2 | NA | 8 |
| 113 | 2 | 2 | 0 | 1 | 1 | 2 | 2 | NA | 10 |
| 45 | 2 | 0 | 1 | 2 | 2 | 2 | 2 | NA | 11 |
| 83 | 2 | 0 | 0 | 2 | 2 | 2 | 2 | NA | 10 |
| 46 | 2 | 2 | 0 | 0 | 0 | 2 | 2 | NA | 8 |
| 94 | 2 | 0 | 0 | 1 | 1 | 2 | 2 | NA | 8 |
| 47 | 2 | 1 | 0 | 0 | 0 | 2 | 0 | NA | 5 |
| 48 | 2 | 0 | 0 | 0 | 0 | 2 | 0 | NA | 4 |
| 49 | 2 | 2 | 0 | 0 | 0 | 2 | 0 | NA | 6 |
| 114 | 2 | 2 | 0 | 0 | 0 | 2 | 2 | NA | 8 |
| 50 | 2 | 2 | 0 | 0 | 0 | 2 | 2 | NA | 8 |
| 95 | 2 | 0 | 0 | 2 | 2 | 2 | 2 | NA | 10 |
| 51 | 2 | 1 | 2 | 0 | 0 | 2 | 2 | NA | 9 |
| 96 | 2 | 0 | 0 | 1 | 1 | 2 | 2 | NA | 8 |
| 121 | 2 | 1 | 0 | 0 | 0 | 2 | 2 | NA | 7 |
| 52 | 2 | 0 | 0 | 2 | 2 | 2 | 2 | NA | 10 |
| 97 | 2 | 2 | 0 | 0 | 0 | 2 | 2 | NA | 8 |
| 98 | 2 | 0 | 0 | 2 | 2 | 2 | 2 | NA | 10 |
| 69 | 2 | 2 | 0 | 0 | 0 | 2 | 2 | NA | 8 |
| 70 | 2 | 2 | 0 | 0 | 0 | 2 | 2 | NA | 8 |
| 115 | 2 | 2 | 0 | 0 | 0 | 2 | 2 | NA | 8 |
| 99 | 2 | 0 | 1 | 1 | 1 | 2 | 2 | NA | 9 |
| 100 | 2 | 2 | 1 | 1 | 1 | 2 | 2 | NA | 11 |
| 101 | 2 | 0 | 0 | 2 | 2 | 2 | 2 | NA | 10 |
| 102 | 2 | 2 | 0 | 1 | 1 | 2 | 2 | NA | 10 |
| 103 | 2 | 0 | 2 | 1 | 1 | 2 | 2 | NA | 10 |
| 71 | 2 | 0 | 2 | 0 | 0 | 2 | 2 | NA | 8 |
| 53 | 2 | 2 | 0 | 1 | 1 | 2 | 2 | NA | 10 |
| 72 | 2 | 0 | 0 | 2 | 2 | 2 | 2 | NA | 10 |
| 81 | 2 | 0 | 0 | 1 | 1 | 2 | 2 | NA | 8 |
| 54 | 2 | 2 | 0 | 1 | 1 | 2 | 2 | NA | 10 |
| 55 | 2 | 2 | 0 | 1 | 1 | 2 | 2 | NA | 10 |
| 56 | 2 | 0 | 1 | 2 | 2 | 2 | 2 | NA | 11 |
| 73 | 2 | 0 | 1 | 2 | 2 | 2 | 2 | NA | 11 |
| 74 | 2 | 1 | 1 | 1 | 1 | 2 | 2 | NA | 10 |
| 75 | 2 | 2 | 1 | 1 | 1 | 2 | 2 | NA | 11 |
| 57 | 2 | 1 | 0 | 1 | 1 | 2 | 2 | NA | 9 |
| 58 | 2 | 0 | 2 | 1 | 1 | 2 | 2 | NA | 10 |
| 76 | 2 | 2 | 0 | 1 | 1 | 2 | 2 | NA | 10 |
| 104 | 2 | 2 | 0 | 1 | 1 | 2 | 2 | NA | 10 |
| 105 | 2 | 2 | 0 | 2 | 2 | 2 | 2 | NA | 12 |
| 106 | 2 | 0 | 2 | 1 | 1 | 2 | 2 | NA | 10 |
| 82 | 2 | 0 | 0 | 2 | 2 | 2 | 2 | NA | 10 |
| 77 | 2 | 1 | 0 | 1 | 1 | 2 | 2 | NA | 9 |
| 116 | 2 | 0 | 2 | 1 | 1 | 2 | 2 | NA | 10 |
| 117 | 2 | 0 | 0 | 2 | 2 | 2 | 2 | NA | 10 |
| 107 | 2 | 1 | 0 | 1 | 1 | 2 | 2 | NA | 9 |
| 108 | 2 | 2 | 2 | 1 | 1 | 2 | 2 | NA | 12 |
| 122 | 2 | 1 | 0 | 0 | 0 | 2 | 2 | NA | 7 |
| 78 | 2 | 0 | 2 | 0 | 0 | 2 | 2 | NA | 8 |
| 118 | 2 | 2 | 0 | 2 | 2 | 2 | 2 | NA | 12 |
| 59 | 2 | 2 | 0 | 0 | 0 | 2 | 2 | NA | 8 |
| 60 | 2 | 0 | 0 | 0 | 0 | 2 | 2 | NA | 6 |
| 119 | 2 | 0 | 2 | 0 | 0 | 2 | 2 | NA | 8 |
| 83 | 2 | 2 | 0 | 0 | 0 | 2 | 2 | NA | 8 |
| 79 | 2 | 0 | 0 | 1 | 1 | 2 | 2 | NA | 8 |
| 84 | 2 | 0 | 1 | 2 | 2 | 2 | 2 | NA | 11 |

*MINORS score is a methodological index for non-randomized studies. The items are scored 0 if not reported; 1 when reported but inadequate; and 2 when reported and adequate. The global ideal score is 16 for non-comparative studies.

As low quality was considered a score of ≤ 8.

As intermediate quality was considered a score of 9-11.

As high quality was considered a score of ≥ 12.

**eTable 3.** Prevalence of organ dysfunctions in adult patients with COVID-19 hospitalized outside the Intensive Care Unit, as reported in included studies (N= 70,822 patients).

| **Ref.** | **N of patients** | **Liver dysfunction, n (%)** | **ARDS, n (%)** | **Coagulopathy, n (%)** | **CNS dysfunction, n (%)** | **Renal dysfunction, n (%)** |
| --- | --- | --- | --- | --- | --- | --- |
| 19 | 62 | 0 (0.0) | 1 (1.6) | 3 (4.8) | 0 (0.0) | 3 (4.8) |
| 23 | 476 | NR | NR | 119 (25.0) | NR | 0 (0.0) |
| 24 | 193 | NR | NR | 10 (5.2) | NR | NR |
| 110 | 123 | 0 (0.0) | 0 (0.0) | 0 (0.0) | 0 (0.0) | 0 (0.0) |
| 85 | 99 | NR | 50 (50.5) | 6 (6.1) | NR | 31 (31.3) |
| 26 | 109 | NR | 84 (77.1) | 16 (14.7) | NR | 12 (11.0) |
| 27 | 1128 | NR | NR | 128 (11.3) | NR | NR |
| 28 | 1476 | NR | NR | 306 (20.7) | NR | NR |
| 61 | 5449 | NR | NR | NR | NR | 1193 (21.9) |
| 29 | 202 | NR | 9 (4.5) | NR | NR | NR |
| 30 | 96 | NR | 25 (26.0) | NR | NR | NR |
| 63 | 72 | NR | 13 (18.1) | NR | NR | 4 (5.6) |
| 31 | 102 | NR | NR | 16 (15.7) | NR | 13 (12.7) |
| 32 | 548 | 24 (4.4) | 207 (37.8) | 154 (28.1) | 17 (3.1) | 146 (26.6) |
| 35 | 116 | NR | NR | NR | NR | 21 (18.1) |
| 65 | 153 | NR | 74 (48.4) | NR | NR | NR |
| 88 | 31 | 0 (0.0) | 31 (100.0) | 4 (12.9) | 0 (0.0) | 9 (29.0) |
| 37 | 154 | 15 (9.7) | 76 (49.4) | 31 (20.1) | 25 (16.2) | 25 (16.2) |
| 89 | 75 | NR | NR | 7 (9.3) | NR | NR |
| 38 | 80 | 1 (1.3) | NR | 11 (13.8) | NR | 2 (2.5) |
| 90 | 200 | 0 (0.0) | 72 (36.0) | NR | NR | 50 (25.0) |
| 39 | 69 | 0 (0.0) | NR | 4 (5.8) | NR | 1 (1.4) |
| 40 | 312 | NR | 25 (8.0) | 51 (16.3) | NR | 5 (1.6) |
| 41 | 13 | NR | 11 (84.6) | 1 (7.7) | NR | 2 (15.4) |
| 91 | 317 | NR | 199 (62.8) | 114 (36.0) | 29 (9.1) | 56 (17.7) |
| 80 | 20133 | NR | NR | NR | 4171 (20.7) | NR |
| 67 | 33 | NR | NR | NR | NR | 22 (66.7) |
| 120 | 200 | NR | 42 (21.0) | NR | NR | NR |
| 42 | 1000 | 64 (6.4) | 140 (14.0) | 99 (9.9) | 10 (1.0) | 116 (11.6) |
| 43 | 304 | NR | NR | NR | 8 (2.6) | NR |
| 112 | 205 | 34 (16.6) | 23 (11.2) | 25 (12.2) | NR | 12 (5.9) |
| 113 | 110 | NR | 18 (16.4) | NR | NR | NR |
| 46 | 703 | 60 (8.5) | NR | 119 (16.9) | NR | 80 (11.4) |
| 94 | 181 | NR | 130 (71.8) | NR | 4 (2.2) | NR |
| 47 | 166 | 55 (33.1) | 75 (45.2) | 16 (9.6) | NR | 19 (11.4) |
| 49 | 62 | NR | NR | NR | NR | NR |
| 114 | 110 | 18 (16.4) | NR | 19 (17.3) | NR | 36 (32.7) |
| 97 | 10021 | NR | NR | NR | NR | 599 (6.0) |
| 69 | 117 | NR | 44 (37.6) | NR | NR | NR |
| 70 | 2512 | NR | 1107 (44.1) | NR | 302 (12.0) | NR |
| 100 | 158 | 41 (25.9) | 34 (21.5) | 35 (22.2) | NR | 15 (9.5) |
| 102 | 8910 | NR | 1834 (20.6) | NR | NR | NR |
| 54 | 187 | 19 (10.2) | 46 (24.6) | 42 (22.5) | NR | 18 (9.6) |
| 55 | 5771 | 358 (6.2) | 577 (10.0) | 28 (0.5) | NR | 115 (2.0) |
| 74 | 156 | NR | 43 (27.6) | 26 (16.7) | NR | 13 (8.3) |
| 75 | 109 | NR | NR | 27 (24.8) | NR | 55 (50.5) |
| 57 | 105 | 1 (1.0) | 11 (10.5) | 26 (24.8) | NR | 11 (10.5) |
| 76 | 1305 | 0 (.0) | 281 (21.5) | NR | 97 (7.4) | 76 (5.8) |
| 104 | 123 | NR | 85 (69.1) | 31 (25.2) | NR | 31 (25.2) |
| 105 | 263 | NR | NR | 33 (12.5) | NR | NR |
| 77 | 3345 | NR | NR | NR | NR | 1903 (56.9) |
| 107 | 2878 | NR | 175 (6.1) | NR | 193 (6.7) | NR |

Abbreviations: ARDS, acute respiratory distress syndrome; NR, not reported.

**eTable 4.** Prevalence of organ dysfunctions in adult patients with COVID-19 in the Intensive Care Unit, as reported in included studies (N= 4,347 patients).

| **Ref.** | **N of patients** | **Liver dysfunction, n (%)** | **ARDS, n (%)** | **Coagulopathy, n (%)** | **Renal dysfunction, n (%)** |
| --- | --- | --- | --- | --- | --- |
| 20 | 52 | 15 (28.8) | 52 (100.0) | NR | 15 (28.8) |
| 109 | 150 | NR | 150 (100.0) | 37 (24.7) | NR |
| 25 | 73 | 33 (45.2) | 55 (75.3) | NR | 13 (17.8) |
| 86 | 35 | NR | 35 (100.0) | 0 (0.0) | 9 (25.7) |
| 111 | 117 | 29 (24.8) | 117 (100.0) | 0 (0.0) | 0 (0.0) |
| 87 | 46 | 4 (8.7) | 46 (100.0) | 3 (6.5) | 10 (21.7) |
| 33 | 239 | 191 (79.9) | 164 (68.6) | 150 (62.8) | 119 (49.8) |
| 36 | 60 | 41 (68.3) | 16 (26.7) | 8 (13.3) | NR |
| 88 | 31 | 0 (0.0) | 31 (100.0) | 4 (12.9) | 9 (29.0) |
| 37 | 154 | 15 99.7) | 76 (49.4) | 31 (20.1) | 25 (16.2) |
| 89 | 75 | NR | NR | 7 (9.3) | NR |
| 41 | 13 | NR | 11 (84.6) | 1 (7.7) | 2 (15.4) |
| 66 | 1150 | 0 (0.0) | 222 (19.3) | 64 (5.6) | 128 (11.1) |
| 44 | 41 | NR | 41 (100.0) | 5 (12.2) | 3 (7.3) |
| 45 | 226 | 42 (18.6) | 161 (71.2) | 66 (29.2) | 70 (31.0) |
| 93 | 80 | NR | 80 (100.0) | 13 (16.3) | 20 (25.0) |
| 48 | 33 | 14 (42.4) | 33 (100.0) | 8 (24.2) | 8 (24.2) |
| 95 | 40 | NR | 40 (100.0) | 0 (0.0) | NR |
| 96 | 90 | 23 (25.6) | 68 (75.6) | 23 (25.6) | NR |
| 52 | 51 | 4 (7.8) | NR | NR | 5 (9.8) |
| 98 | 379 | NR | NR | 40 (10.6) | 193 (50.9) |
| 101 | 92 | NR | NR | 4 (4.3) | NR |
| 103 | 30 | NR | NR | NR | 15 (50.0) |
| 53 | 244 | 0 (0.0) | 153 (62.7) | 11 (4.5) | NR |
| 81 | 187 | NR | 187 (100.0) | NR | NR |
| 56 | 59 | NR | NR | NR | 23 (39.0) |
| 73 | 217 | NR | 165 (76.0) | NR | NR |
| 58 | 103 | NR | 99 (96.1) | 26 (25.2) | 6 (5.8) |
| 106 | 56 | NR | 56 (100.0) | NR | NR |
| 82 | 39 | NR | 39 (100.0) | NR | NR |
| 107 | 185 | NR | 27 (14.6) | NR | NR |

Abbreviations: ARDS, acute respiratory distress syndrome; NR, not reported.

**eTable 5.** Prevalence of specific organ dysfunctions in children hospitalized with COVID-19, as reported in included studies (N= 303).

| **Ref.** | **N of patients** | **Shock, n (%)** | **Liver dysfunction, n (%)** | **ARDS, n (%)** | **Coagulopathy, n (%)** | **CNS dysfunction, n (%)** | **Renal dysfunction, n (%)** |
| --- | --- | --- | --- | --- | --- | --- | --- |
| 118 | 81 | 2 (2.5) | 0 (0.0) | 1 (1.2) | 0 (0.0) | 0 (0.0) | 0 (0.0) |
| 59 | 40 | NR | NR | NR | NR | NR | 0 (0.0) |
| 60 | 34 | 0 (0.0) | NR | 3 (4.6) | NR | NR | NR |
| 79 | 33 | 17 (51.5) | 8 (24.2) | NR | 8 (24.2) | NR | 8 (21.6) |
| 119 | 37 | 1 (2.7) | NR | NR | 15 (40.5) | 2 (5.4) | NR |
| 84 | 78 | 1 (1.3) | 68 (87.2) | NR | NR | 39 (50.0) | NR |

Abbreviations: ARDS, acute respiratory distress syndrome; NR, not reported.

**eTable 6.** Summary of inflammation markers in the included studies.

| **Ref.** | **N.** | **Lymphocyte count (×10^9^/L)** | | **C-Reactive Protein (mg/L)** | | **Procalcitonin (ng/mL)** | | **Ferritin (ng/mL)** | **IL-6 (pg/mL)** |
| --- | --- | --- | --- | --- | --- | --- | --- | --- | --- |
|  |  |  | **↓ n (%)** |  | **↑ n (%)** |  | **↑ n (%)** |  |  |
| 19 | 62 | 1.0 (0.8-1.5) | 26 (42) |  |  | 0.04 (0.03-0.06) | 7 (11) |  |  |
| 21 | 85 | 0.7 ± 0.4 | 66 (76.6) | 107.26 ± 117.22 | 78 (91.8) | 3.65 ± 13.40 | 19 (22.4) |  |  |
| 23 | 476 | 1.0 (0.7-1.5) | 225 (47.3) | 18.8 (5.2-57.0) | 266 (64.1) | 0.05 (0.02-0.08) |  |  |  |
| 25 | 73 | 0.7 (0.5-0.9) |  | 872 (326-1045) |  | 0.1 (0.0- 0.24) |  |  |  |
| 85 | 99 | 0.9 (0.7-1.3) |  | 650 (210-1330) |  | 0.3 (0.1-0.8) |  | 1392 (745-2733) |  |
| 26 | 109 | 0.6 (0.4-0.9) | 90 (82.6) | 85.7 ± 57.3 | 104 (95.4) | 0.1 (0.1-0.3) | 20 (18.3) |  |  |
| 86 | 35 | 0.7 (0.5-1.2) |  | 106.0 (54.5-262) |  |  |  |  | 26 (11–69) |
| 66 | 257 | 0.8 (0.6-1.2) |  | 158 (92-254) |  | 0.35 (0.17-1.1) |  | 924 (472–1789) |  |
| 45 | 226 | 0.8 (0.5-1.2) | 160 (70.8) |  |  | 0.19 (0.05-1.4) | 82 (37.3) |  |  |
| 95 | 40 |  |  |  |  | 0.52 ± 0.25 |  |  | 108.4 ± 91.1 |
| 29 | 202 | 1.1 (0.8- 1.6) | 66 (32.7) |  | 55 (27.2) |  | 15 (11.4) |  |  |
| 63 | 72 |  |  |  | 36/41 (87.8) |  | 4/47 (8.5) | 824 (453-1643) |  |
| 31 | 102 | 0.9 (0.6-1.2) | 66 (65) | 34.0 (5.8-86.6) | 86 (84) | 0.06 (0.03-0.15) |  |  | 4.7 (2.2-20.3) |
| 118 | 81 | 2.3 (0.4-13.4) | 4 (5) | 2 (0-139) | 13 (16) | 0.08 (0.02-2.22) | 3 (3.7) |  |  |
| 33 | 239 | 0.6 (0.5-0.8) | 219 (91.6) |  |  |  |  |  | 9.1 (6.7-12.0) |
| 34 | 107 | 1.0 (0.7-1.5) |  | 35.8 (5.0-85.1) |  |  |  |  |  |
| 36 | 60 |  | 38 (63.3) |  | 7 (11.7) |  |  |  |  |
| 65 | 153 | 0.9 (0.6-1.2) | 89 (58) |  |  |  |  |  |  |
| 38 | 80 | 0.6 (0.4-1.0) | 26 (32.5) | 6.6 (5.3-12.3) | 62 (77.5) | 1.3 (0.4-2.6) | 1 (1.25) |  |  |
| 90 | 200 | 1.2 (0.9-1.8) |  | 38.1 (13.3-93.5) |  |  |  | 1032 (591-1638) | 41 (21–97) |
| 40 | 312 | 1.1 (0.8-1.4) | 146 (51) | 17.6 (3.8-41.3) | 166 (65) | 0.10 (0.06-0.16) | 11 (8) |  |  |
| 41 | 41 | 0.8 (0.6-1.1) | 15 (37) |  |  | 0.1 (0.1-0.1) | 3 (8) |  |  |
| 91 | 317 |  | 192/281 (68.3) | 79.3 (33.7-132.5) |  |  |  |  | 46.7 (20-97.9) |
| 73 | 217 |  |  | 190 (126-262) |  |  |  |  |  |
| 75 | 109 |  |  | 146 (101-227) |  | 0.23 (0.12-0.92) |  | 579 (339-1057) | 14 (7.5-40.5) |
| 57 | 105 | 1.1 (0.7-1.4) | 33 (31.4) | 33.6 (9.4-56.9) | 83 (79.0) |  |  |  |  |
| 76 | 1,305 | 0.95 (0.32-0.61) |  | 115 (64.7-180.8) |  | 0.18 (0.08-0.48) |  |  |  |
| 104 | 123 | 0.98 (0.74-1.22) |  | 90.2  (44.7-144.6) |  |  |  |  |  |
| 79 | 33 | 1.1 (0.6-1.3) |  | 250 (156-302) |  | 5.4 (1.8-16.7) |  | 568 (340-954) | 200 (56.4-330) |
| 106 | 56 | 0.9 (0.7-1.2) |  | 152 (116-210) |  |  |  | 1376 (1023-6927) | 60 (36.4-126) |
| 84 | 78 | 0.7 (0.4-1.1) |  | 264 (192–316) |  |  |  | 1042 (538-1746) |  |
| 107 | 1240 | 1.3 ± 3.6 |  | 91 ± 77 |  |  |  | 1235 ± 2483 |  |
| 82 | 39 | 0.76 ± 0.4 |  | 305 ± 101 |  |  |  | 987 (552-1425) |  |
| 77 |  |  |  | 108 (48-191) |  | 0.2 (0.1-0.7) |  | 762.5 (384-1420) |  |
| 81 | 187 | 0.8 (0.5-1.1) |  | 202 (128-294) |  |  |  | 1126 (495-1880) |  |
| 101 | 92 | 0.8 (0.6-1.1) |  | 175 (131-232) |  | 0.9 (0.3-2.2) |  |  |  |
| 100 | 158 |  | 127/145  (87.6) |  | 67/147  (45.6) |  | 12/83 (14.5) |  |  |
| 99 | 196 |  |  | 81 (40-132) |  |  |  | 762 (452-1424) | 36.9 (22-64) |
| 69 | 117 | 1.53 ± 3.03 |  | 97.6 ± 62.7 |  | 0.59 ± 1.49 |  | 1173.8 ± 1623.2 |  |
| 52 | 51 | 0.7 (0.4-0.8) |  | 53.0 (19.7-84.7) |  | 0.15 (0.05-0.34) |  |  |  |

Data are presented as mean ± standard deviation or median (quartile 1-3)

↓↑: number of patients (%) with decrease or increase of the respective marker

Abbreviation: N number of patients.

**eTable 7.** Summary of coagulation markers in the included studies.

| **Ref.** | **N.** | **d-dimers (mg/L)** | | **Fibrinogen (g/L)** | **aPTT (s)** | **PT (s)** |
| --- | --- | --- | --- | --- | --- | --- |
|  |  |  | **↑ n (%)** |  |  |  |
| 19 | 62 | 0.2 (0.2-0.5) |  |  |  |  |
| 109 | 150 | 2.27 (1.16-20) |  | 6.99 (6.08-7.73) |  |  |
| 21 | 85 | 0.0052 ± 0.0047 | 56 (55.9) | 6.32 ± 18.35 | 39.2 ± 9.3 |  |
| 23 | 476 | 0.58 (0.35-1.48) |  | 4.4 (3.65–5.41) |  |  |
| 25 | 73 | 0.6 (0.4-3.4) |  |  | 36.2 (30.4-40.8) | 14.2 (12.6- 15.6) |
| 85 | 99 | 0.58 (0.33-0.99) |  |  |  |  |
| 26 | 109 | 1.4 (0.6-4.8) | 85 (78.0) |  | 34.3 ± 10.7 | 12.9 (11.6-15.4) |
| 86 | 35 | 4.01 (1.41-14.40) |  |  |  |  |
| 66 | 257 | 1.6 (0.9-3.5) |  |  |  | 14.7 (14.0-15.8) |
| 45 | 226 | 3 (1.2-7.1) | 189 (83.6) |  | 32.3 (26.1-42.1) | 13 (11.6-14.7) |
| 95 | 40 | 1.56 ±1.09 |  | 8.96 ± 1.10 | 32.2 ± 2.9 | 65.1 ± 9.8 |
| 29 | 202 |  |  |  |  | 12.8 (12.0- 13.4) |
| 63 | 72 |  | 4/47 (8.5) |  |  |  |
| 31 | 102 | 0.8 (0.5-1.7) | 74 (72.5) |  |  | 14.2 (13.7-14.8) |
| 118 | 81 | 0.22 (0.15-1.57) | 10 (12.2) |  |  | 9.3 (7.8-16.9) |
| 33 | 239 |  |  |  | 29.3 ± 9.0 | 11.9 ± 1.7 |
| 34 | 107 | 0.76 (0.36-2.14) |  | 4.54 ± 1.57 | 28.1 (26.1-30.6) | 12 (11.3-13) |
| 37 | 154 |  | 91 (59.1) |  |  |  |
| 89 |  | 1.1 (0.7-2.3) | 185 (93.4) |  |  |  |
| 38 | 80 | 0.9 (0.4-2.4) | 3 (3.8) |  | 18.6 (17.2-28.6) | 10.8 (9.3-13.2) |
| 90 | 200 | 1.2 (0.5-2.9) |  |  |  |  |
| 40 | 312 | 0.44 (0.22–0.97) | 86 (28) | 3.92 (3.05-4.93) | 36.8 (32.7-41.1) | 12.9 (12.0-13.8) |
| 41 | 41 | 0.5 (0.3-1.3) |  |  | 27.0 (24.2-34.1) | 11.1 (10.1-12.4) |
| 91 | 317 | 1.05 (0.63-1.57) |  |  |  |  |
| 54 | 187 | 0.43 (0.19-2.66) |  |  | 32.0 (30.1-35.0) | 12.8 (12.0-14.0) |
| 73 | 217 | 1.73 (0.93-6.95) |  |  |  |  |
| 75 | 109 | 0.51 (0.32-0.97) |  | 5.35 (4.35-6.51) |  |  |
| 57 | 105 | 0.6 (0.3-1.9) | 40 (38.1) |  | 33.9 (30.0-37.9) | 12.5 (11.4-13.1) |
| 76 | 1,305 | 1.15 (0.68-2.53) |  |  | 33.1 (30.2-37.5) | 13.2 (12.4-14.4) |
| 79 | 33 | 3.7 (2.4-5.1) |  | 6.27 (4.55-7.82) |  |  |
| 106 | 56 | 1.41 (0.78-2.29) |  |  |  |  |
| 84 | 78 | 4.03 (2.35-7.42) |  |  |  |  |
| 107 | 1240 | 1.64 ± 4.21 |  | 6.1 ± 1.7 |  |  |
| 82 | 39 | 6.44 ± 10.43 |  | 6.6 ± 1.9 | 38.8 ± 13.1 | 14.1 ± 2.1 |
| 77 |  | 1.7 (0.9-3.8) |  | 6.49 ± 2.0 |  |  |
| 81 | 187 | 2.59 (0.95-10.0) |  | 7.0 (6.0-10.0) |  | 12.4 (11.0–14.9) |
| 101 | 92 |  |  | 7.7 (6.1-8.8) |  |  |
| 100 | 158 |  | 68/121 (56.2) |  |  |  |
| 99 | 196 | 1.13 (0.67-1.73) |  |  |  |  |
| 69 | 117 | 2.23 ± 3.29 |  |  | 36.06 ± 11.50 | 16.00 ± 6.16 |
| 52 | 51 |  |  |  | 33.6 (29.3-37.9) | 12.8 ± 1.7 |

Data are presented as mean ± standard deviation or median (quartile 1-3)

↓↑: number of patients (%) with decrease or increase of the respective marker

Abbreviations: N number of patients; PT prothrombin time; aPTT activated partial thromboplastin time.
